# Familial medullary thyroid carcinoma secondary to an *SLC30A9* intragenic deletion and translation reinitiation

**DOI:** 10.64898/2026.02.26.26346165

**Authors:** Donato Iacovazzo, Federica Begalli, Oniz Suleyman, Márton Doleschall, Maria Alevizaki, Kevin E. Ashelford, Sherine Awad, Anne Barlier, Sayka Barry, Caroline Brain, Claudia P. Cabrera, Frédéric Castinetti, Sabrina Chiloiro, Kevin Colclough, Alexandra Csábi, Maralyn R. Druce, Pinaki Dutta, Jawid M. Fatih, William D. Foulkes, Mira Gandhi, Christopher M. Grochowski, Charlotte L. Hall, Barbara Jarząb, Kathleen O. Klein, Jolanta Krajewska, Tom R. Kurzawinski, Sebastiaan Lamers, Francesca Lugli, Kesson Magid, Rebecca Margraf, Carmen S. Martin, Jes S. Mathiesen, Radu Mihai, Patrick J. Morrison, Monika Mozere, Małgorzata Oczko-Wojciechowska, Martina Owens, Luka Ozretić, Attila Patócs, Serena Piacentini, Jaya Punetha, Ashutosh Rai, Pauline Romanet, Suvi Savola, Stefan Schoenfelder, David M. Scott-Coombes, Horia Stanescu, Mehmet Tekman, Laura E. Thomas, Miklós Tóth, Steven W. Wingett, Michael Witcher, Claudia M.B. Carvalho, Martin Franke, Robert Kleta, James R. Lupski, Julian R. Sampson, Laura De Marinis, Márta Korbonits

**Author notes:** these authors contributed equally (Federica Begalli, Oniz Suleyman).

## Abstract

While most individuals with familial medullary thyroid carcinoma (fMTC) carry *RET* mutations, in some instances the causative mutations remain unknown. We studied two related families with *RET*-negative fMTC in 21 affected individuals through linkage analysis, exome/genome sequencing, and high-density array comparative genomic hybridization. We identified a novel heterozygous 40kb intragenic *SLC30A9* deletion which segregated with the disease in all affected individuals. The mutant transcript escaped nonsense-mediated decay and resulted in the production of N-terminally truncated proteins via translation reinitiation from in-frame AUG codons located downstream of the deletion. These proteins localized to the endoplasmic reticulum suggesting protein misfolding. Introducing the deletion in an MTC cell line resulted in significant upregulation of genes involved in the unfolded protein response, ER-associated degradation, and the PI3K-Akt pathway. Furthermore, the deletion was associated with increased cell proliferation, migration, and clonogenic capacity, supporting an oncogenic role. These findings expand the genetic background of fMTC beyond *RET* mutations and implicate translation reinitiation in the etiology of cancer susceptibility syndromes secondary to structural genomic variants.

## Introduction

Medullary thyroid carcinoma (MTC) is a neuroendocrine tumor originating from the calcitonin-expressing C cells of the thyroid gland with an estimated annual incidence ranging from 0.14 to 0.21/100,000 population in the United States^1^. MTC accounts for around 2% of thyroid cancers and 8-13% of thyroid cancer-related mortality^2,3^. MTC occurs as sporadic disease in approximately 75% of cases, while in the remaining cases it presents in a familial setting as a spectrum of autosomal dominant conditions including multiple endocrine neoplasia type 2A (MEN2A), multiple endocrine neoplasia type 2B (MEN2B), and familial MTC (fMTC). The latter is characterized by the occurrence of MTC without other associated manifestations^4^. Familial forms of MTC are generally caused by activating mutations in the *RET* proto-oncogene (OMIM 164761). While virtually all cases of MEN2A and MEN2B are caused by *RET* mutations, a subset (<5%) of families with fMTC do not carry pathogenic *RET* mutations, and the causative genetic mutations in these families remain to be identified. The exact prevalence of *RET* mutation-negative fMTC is difficult to assess due to the rarity of the phenotype. In a large cohort of 250 Italian families with hereditary MTC, six families (2.4%) did not carry pathogenic *RET* mutations^5^. In a Greek cohort, 3.2% of 313 patients with heritable MTC did not carry *RET* mutations^6^, and a further six families with *RET* mutation-negative fMTC were reported in a Spanish study^7^. A large kindred with 19 cases of *RET* mutation-negative fMTC has been recently reported^8^. Collectively, these reports suggest that inherited pathogenic variants in genes other than *RET* are involved in the etiology of fMTC.

We have studied two large, related families with *RET* mutation-negative fMTC, using linkage analysis, next-generation sequencing, and high-density array comparative genomic hybridization (HD-aCGH). We identified a novel intragenic deletion in the *SLC30A9* gene which was found to fully segregate with the phenotype and carrier status over multiple generations. The intragenic deletion resulted in translation reinitiation from in-frame AUG codons located downstream of the deletion and production of N-terminally truncated SLC30A9 proteins which localized within the endoplasmic reticulum (ER), suggesting protein misfolding. Introducing the deletion in an MTC cell line resulted in the enrichment of genes involving the unfolded protein response and the PI3K-Akt pathway, as well as increased cell proliferation, migration, and clonogenic capacity, implicating this unique deletion in the pathogenesis of fMTC.

## Results

### Clinical data

The index family (family 1; Figure 1A) had a history of thyroid cancer in 13 family members across four generations. MTC was confirmed in 12 individuals, while one subject (II/3) had thyroid cancer without available histology to confirm MTC. No family members showed signs of catecholamine excess or primary hyperparathyroidism, supporting a diagnosis of fMTC with an autosomal dominant inheritance pattern. There was no history of cutaneous lichen amyloidosis or Hirschsprung disease. X-linked inheritance was excluded based on multiple instances of father-to-son transmission, and Sanger sequencing of the entire coding region of *RET* revealed no pathogenic mutations. The second family (family 2; Figure 1B) showed a similar history with four confirmed MTC cases and at least two other subjects (I/1 and II/2) known to have had surgery for thyroid cancer. Similar to family 1, there was no history of pheochromocytoma. One subject (family 2: III/2) was diagnosed with primary hyperparathyroidism secondary to a single parathyroid adenoma. No pathogenic mutations were identified in this family following sequencing of the entire coding region of *RET*. As subsequent genetic analyses confirmed relatedness between the two kindreds, they will be referred to as index families.

**Figure 1.**
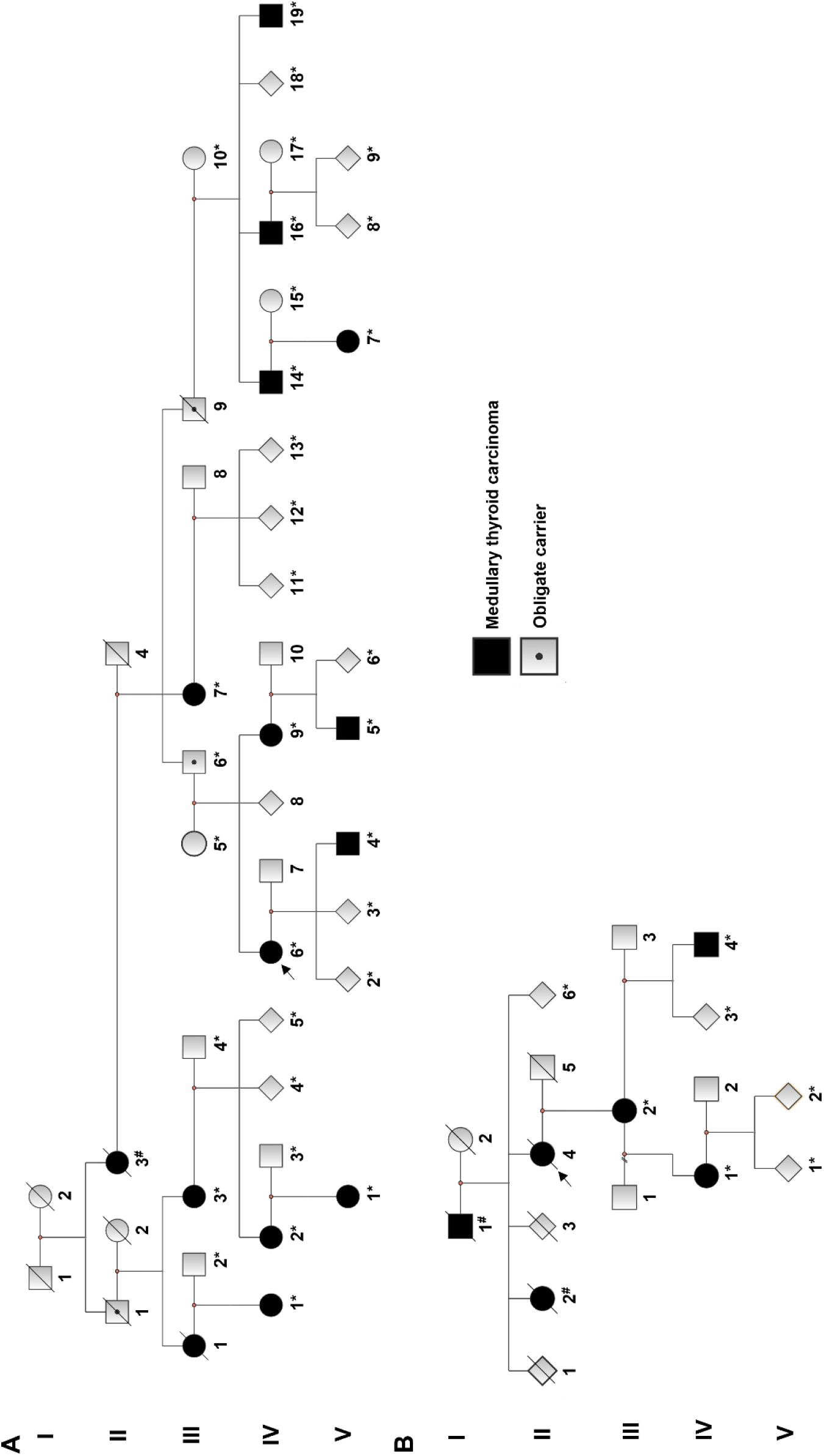
Family trees of the two index families with *RET* mutation-negative fMTC. Patients diagnosed with MTC (or thyroid cancer for those subjects^#^ in the older generations where a diagnosis of MTC could not be confirmed) are highlighted in black. Obligate carriers are highlighted with a black dot. The asterisks mark subjects for whom a DNA sample was available. Probands are highlighted by arrows. To improve readability, a simplified version of the family trees is shown.

Out of the 19 affected individuals in the two families, six presented clinically with a thyroid nodule or metastatic disease, while the remaining 13 subjects were diagnosed prospectively following clinical screening. Mean age at diagnosis was 22.3±9.7 years (range 7-41 years). Seventeen subjects underwent surgery (total thyroidectomy +/-lymph node dissection). Treatment outcomes were variable, with four patients having persistent or recurrent disease. Lymph node metastases were confirmed pathologically in four subjects (out of 10 with available data). Two patients were diagnosed with papillary thyroid carcinoma. In one case (family 1: IV/18), a papillary thyroid microcarcinoma [stage pT1a(m)] – in the absence of concomitant MTC – was diagnosed following screening neck ultrasound. In the second case (family 1: V/4), an incidental focus of papillary thyroid microcarcinoma (stage pT1a) was discovered following total thyroidectomy for MTC. The relatively high frequency of papillary thyroid carcinoma in the general population suggests that this represents a coincidental association.

The prospectively identified affected subjects had an overall good prognosis, with 11 out of 13 patients having long-term disease remission following surgical treatment (mean follow-up 21.7±11.3 years; range 6-36 years). Overall, at least two individuals were known to have died from metastatic MTC in the index families. One other individual (family 1: II/3) was known to have died from metastatic thyroid cancer, although a diagnosis of MTC could not be confirmed. Clinical details are shown in Supplementary Table 1. Notably, the age at diagnosis in seven additional non-index families with either *RET* mutation-negative fMTC (n=5) or fMTC-like phenotype (n=2; Supplementary Figure 1) was significantly higher compared to the index families and aligned with sporadic MTC (46.3±9.3 years, range 37-64 years; *P* <0.0001).

### Histology and tumor-based studies

Histology from nine available cases from the index families showed multifocal tumors composed of sheets and nests of epithelioid to focally spindled cells with uniform nuclei and characteristic “neuroendocrine” type finely stippled chromatin (Figure 2A-B), stromal amyloid deposition (Figure 2C), and diffuse positivity for calcitonin with immunohistochemistry (Figure 2D), alongside chromogranin and synaptophysin. The mitotic count was <2 mitoses/2mm^2^, the Ki-67 proliferation index was low (<3% in all cases; Figure 2F), and these was no necrosis, in keeping with low-grade MTC^9^. The adjacent thyroid parenchyma showed foci of C cell hyperplasia (Figure 2E).

**Figure 2.**
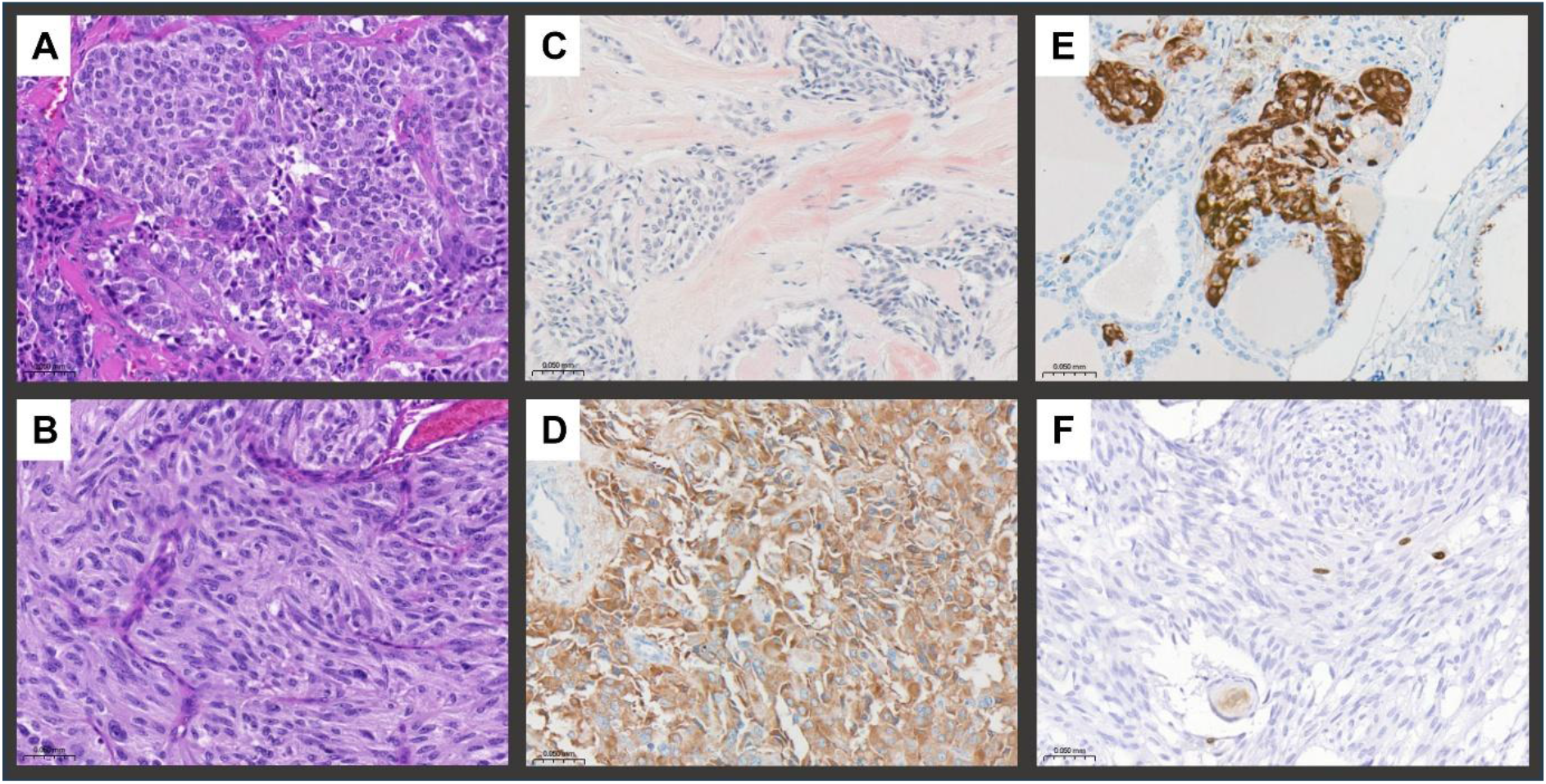
Histopathology and representative immunostaining of MTC tumor samples from the index families. The tumors showed nested architecture and were composed of epithelioid (A) to spindled (B) cells with characteristic speckled ‘neuroendocrine type’ chromatin. There was deposition of stromal eosinophilic acellular material showing typical salmon pink coloration on Congo Red stain in keeping with amyloid (C). The tumor cells were diffusely positive for calcitonin (D). The surrounding thyroid parenchyma showed areas of nodular C cell hyperplasia (E). The Ki-67 proliferation index was low (<3%; F). Magnification x20.

No somatic hotspot *RET* (codons 634, 883, and 918) or *RAS* (*HRAS*, *KRAS,* and *NRAS* – codons 12-13 and 61) mutations were detected in tumor DNA with Sanger sequencing, further indicating that tumorigenesis in these families was independent from driving mutations commonly identified in sporadic MTC.

An OncoScan CNV assay highlighted chromosomal changes, many of which were previously described in MTC^10,11^, with no evidence of loss-of-heterozygosity at the 4p13 locus (Supplementary Figure 2). Furthermore, there were no somatic gains or losses involving the *RET* gene and no somatic mutations in nine cancer-related genes (*BRAF*, *EGFR*, *IDH1*, *IDH2*, *KRAS*, *NRAS*, *PIK3CA*, *PTEN,* and *TP53*).

### Identification of a novel *SLC30A9* intragenic deletion

Single nucleotide polymorphism (SNP) genotyping showed an excess of relatedness between the two index families: the calculated PI_HAT (Proportion Identity-by-descent Hat) score^12^ for subjects III/7 (family 1) and III/2 (family 2) was 0.06, while the expected score of 0 was obtained when testing unrelated subjects. Subsequently, linkage analysis was performed with the aim of determining linked regions among affected subjects. Non-parametric linkage analysis highlighted a linkage region with maximum logarithm of the odds (LOD) score of 3.93 on chromosome 4 (27,663,167-54,931,808; hg19) (Figure 3A-B). Haplotype reconstruction revealed two shared regions across all affected subjects of about 2.5Mb (from rs1898889 to rs1948314) and 1Mb (from rs10049629 to rs11733919), including 11 protein-coding genes (Supplementary Table 2).

**Figure 3.**
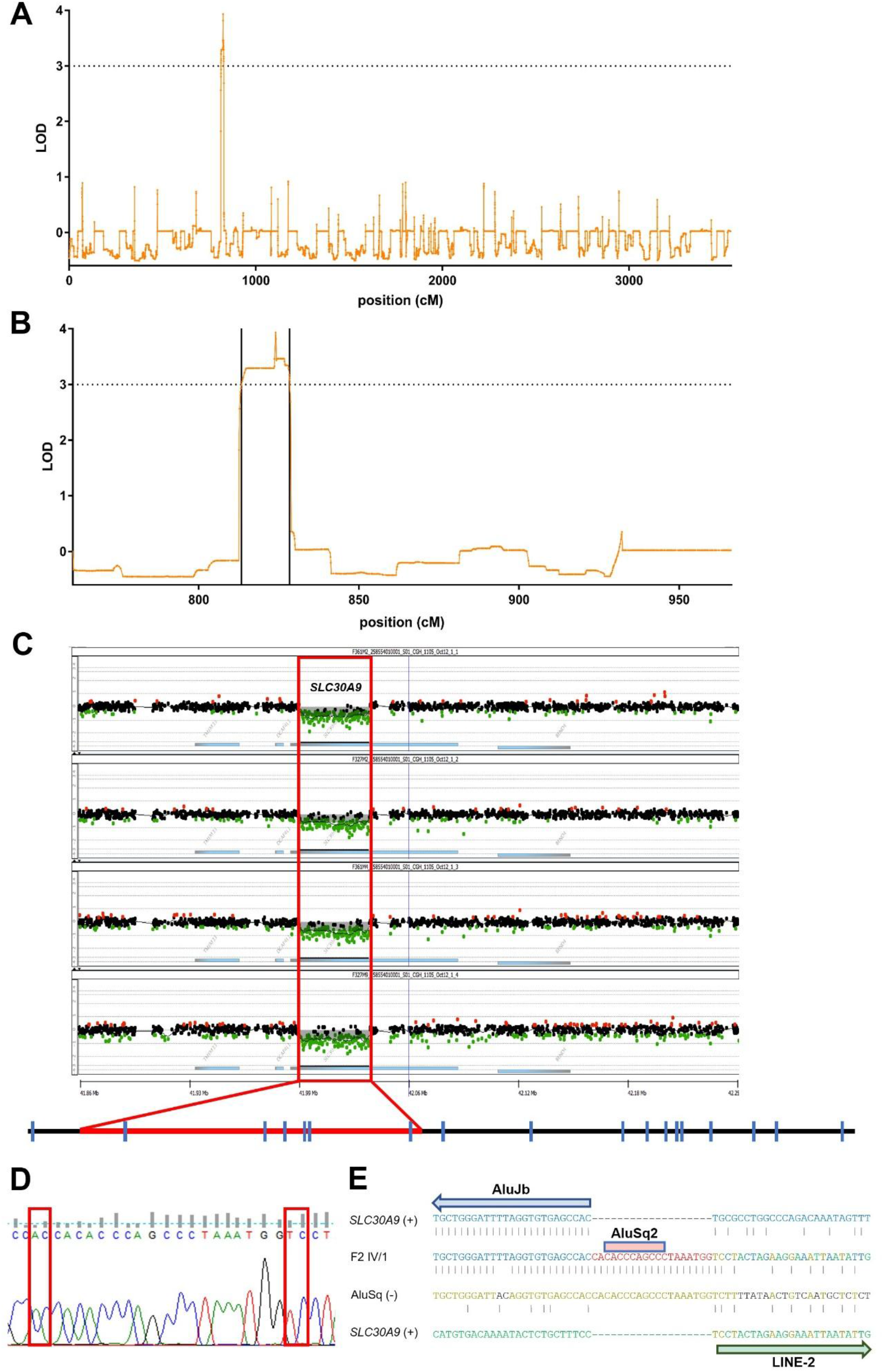
Linkage analysis and identification of an *SLC30A9* intragenic deletion in the two *RET* mutation-negative fMTC index families. A. Non-parametric linkage based on 21,115 single nucleotide polymorphisms across the index families, genome-wide results. B. Non-parametric linkage, chromosome 4 view. The LOD score is represented on the y axis, while genomic position in centimorgan (cM) is visualized on the x axis. A region of approximately 23Mb with maximum LOD score of 3.9 is seen in chromosome 4. C. Top: log-2 ratio of the custom high-density CGH array showing a heterozygous intragenic deletion (red box) in the *SLC30A9* gene in all four tested subjects. Bottom: scheme of the *SLC30A9* gene (exons are shown as blue bars) with the intragenic deletion encompassing exons 2 to 7 highlighted in red. D. The deletion was confirmed by PCR and Sanger sequencing in all affected subjects. Breakpoint junctions are highlighted by red rectangles. E. Genomic architecture at the deletion site. Top and bottom lines show the *SLC30A9* genomic sequence upstream (blue letters) and downstream (green letters) of the deletion, respectively. The second line shows the sequence of the allele carrying the deletion, with the 19bp insertion at the breakpoint junction in red. The third line shows the genomic sequence around chr4:41,996,985 (hg19) where the insertion is likely copied from. Repetitive sequences (Alu and LINE-2 elements) are observed, suggesting microhomology-mediated break-induced replication as the causative mechanism. Microhomologies are highlighted by black lines and letters of the same color.

Having defined the genomic region likely encompassing the causative mutation, exome sequencing (family 1: IV/1, IV/6, and V/7; family 2: III/2) and genome sequencing (family 1: IV/16 and family 2: IV/1) were performed. We assumed a rare autosomal dominant model of inheritance to filter novel heterozygous variants (not previously reported in the GnomAD, ExAC, dbSNP, and 1,000 Genomes databases) annotated as missense, nonsense, frameshift, non-frameshift deletions or insertions, and splice site variants. No variants were identified in the shared haplotypes despite high coverage of all coding regions; similarly, no shared variants were identified in this region after including previously reported (minor allele frequency [MAF] <0.05%) or synonymous variants. Three novel heterozygous missense variants were identified genome-wide across all tested individuals, although *in silico* prediction tools strongly favored them to be benign (Supplementary Table 3). No variants were identified in the *ESR2* gene, encoding estrogen receptor β, which had been previously implicated in the pathogenesis of MTC in a single kindred with an fMTC-like phenotype^13^. Similarly, there were no pathogenic variants in the *MET* gene^14^. Four common *RET* synonymous variants were identified in the affected subjects, all of which are considered benign according to the guidelines of the American College of Medical Genetics (Supplementary Table 4). Confirming the results of Sanger sequencing, no pathogenic *RET* variants were found.

Analysis of non-coding variants revealed 20 rare (MAF <0.05%) heterozygous variants in the shared haplotypes, including 15 intergenic and five intronic variants (Supplementary Table 5). *In silico* tools did not predict effects on gene splicing for any of the intronic variants identified. Furthermore, none of these variants fell within genomic regions with possible regulatory function, as assessed by the absence of H3K4me1/H3K27ac histone marks or transcription factor binding sites in publicly available datasets.

Subsequently, HD-aCGH with a high density of probes on chromosome 4 (15-70Mb) was used to test DNA from four affected individuals (family 1: IV/2, IV/19 and family 2: III/2, IV/1) with the aim of identifying any shared copy number variants (CNVs). The array showed four CNVs within the shared haplotypes (Supplementary Table 6). Only one of these was found in all four tested subjects: a novel heterozygous intragenic deletion encompassing exons 2 to 7 of the *SLC30A9* gene (4p13), encoding a member of the SLC30 family of zinc transporters (Figure 3C). Interrogation of genome sequencing data revealed split reads at the deletion site, supporting the presence of a deletion. Sequencing of the breakpoints confirmed a heterozygous deletion of 40,348bp (chr4:41,998,016-42,038,363; hg19). A 19bp insertion was seen at the deletion site, which mapped to the reverse complement of a sequence around 1kb upstream of the deletion site; microhomology was observed in this region, suggesting microhomology-mediated break-induced replication (MMBIR) as the likely mechanism to have caused this CNV (Figure 3D-E)^15^. Importantly, the deletion fell within the shared haplotypes and was identified by Sanger sequencing to fully segregate with the phenotype – i.e. all affected subjects and obligate carriers with available DNA from the two index families were found to carry the deletion (Supplementary Figure 3A). RT-PCR of peripheral blood-derived cDNA from affected subjects showed an aberrant transcript (Supplementary Figure 3B) comprising exons 1 and 8, in keeping with the deletion of exons 2 to 7. Importantly, no other cases of deletions limited to this region are reported in the DGV, DECIPHER, ClinVar, and GnomAD databases.

By way of prospective genetic screening of at-risk subjects, two additional carriers of the deletion were identified (family 1: III/7 and V/1). Clinical screening in these subjects showed elevated calcitonin levels (III/7: 625.8 pg/ml, normal value <9.8; V/1: 14.3 pg/ml, normal value <11.5). Both patients underwent surgery and histology confirmed the presence of multifocal MTC (with associated lymph node metastases in one case; Supplementary Table 1).

No pathogenic *SLC30A9* single nucleotide variants or small insertions/deletions were identified with targeted sequencing in the seven non-index families with *RET* mutation-negative fMTC or fMTC-like phenotype as well as in a cohort of 108 patients (85 leukocyte-derived and 23 tumor-derived DNA samples) with *RET* germline mutation-negative sporadic MTC. Testing by means of multiplex ligation-dependent probe amplification (MLPA) and HD-aCGH in the seven families and 62 patients with sporadic MTC did not detect any CNVs involving the *SLC30A9* gene.

The expression of SLC30A9 was investigated by immunohistochemistry using a C-terminus directed antibody in seven tumor samples from the index families and in a group of 10 sporadic MTC controls. SLC30A9 expression was found to be reduced in the tumor samples from patients carrying the *SLC30A9* deletion compared to the sporadic MTC controls (mean H-score 100.3±47.4 vs 164.6±18.26, *P* value 0.0013) (Supplementary Figure 4).

### The deletion results in translation reinitiation and truncated SLC30A9 forms with oncogenic activity

Since the mutation is predicted to result in a frameshift with a premature stop codon in exon 8 – p.(Glu37Alafs*11) - we first evaluated the effects of *SLC30A9* knockdown in the MTC TT cell line^16^. Following short hairpin RNA-mediated *SLC30A9* silencing (Supplementary Figure 5A-C), no changes in cell viability were found compared to the non-targeting control (Supplementary Figure 5D). Similarly, no consistent differences were observed in terms of ERK and AKT activation (Supplementary Figure 6). Given the known association of biallelic inactivating *SLC30A9* variants with Birk-Landau-Perez syndrome^17–22^ these findings suggested that mechanisms other than *SLC30A9* loss-of-function are responsible for the MTC phenotype.

To assess broader regulatory effects of the deletion, we performed RNA-Seq in patient-derived EBV-transformed lymphoblastoid cells and skin fibroblasts. Among the eight protein-coding genes within +/-1Mb of the deletion, only *SHISA3*, located ∼400kb away, was differentially expressed in patient-derived cells when compared with healthy controls (upregulated in both the lymphoblastoid cells and skin fibroblasts; Supplementary Table 7). To further investigate any regulatory effects, Hi-C was performed in TT cells and the results were compared to Hi-C data from NHEK cells of epidermal origin^23^. In both cell types, we observed a similar pattern of chromatin interactions and organization of topologically associating domains (TADs) across a 1.6Mb region surrounding *SLC30A9*. Neighboring gene loci were separated by distinct TADs, including *LIMCH1* and *PHOX2B* on the telomeric side, and *SHISA3* on the centromeric side of *SLC30A9* (Supplementary Figure 7A). Notably, the region containing the deletion did not appear to participate in long-range interactions in either cell type. Consistent with this, we found no evidence for the binding of architectural DNA-binding factors across cell types, such as CTCF or cohesin complex components, nor binding sites for other transcription factors (ENCODE data^24^; data not shown). To investigate high-resolution changes in chromatin interactions induced by the deletion, we performed UMI-4C^25^, targeting the promoters of *SLC30A9*, *BEND4*, and *SHISA3* in skin-derived fibroblasts from an affected patient and a healthy control (Supplementary Figure 7B). Chromatin interactions from the *SLC30A9* and *BEND4* promoters overlapped and were confined by adjacent TAD boundaries, as predicted by Hi-C data. In contrast, chromatin interactions from the SHISA3 promoter were spatially distinct and restricted to its Hi-C-predicted TAD. Differential analysis of chromatin interactions from the *SLC30A9* promoter showed a loss of contacts within the deleted region and a gain of interactions within the gene body, consistent with the centromeric position of the deletion. The chromatin interactions of other genes remained unchanged. Confirming these findings, no significant differences were seen when the expression of SHISA3 was investigated by immunohistochemistry in seven tumor samples from the index families compared to 10 sporadic MTC controls (mean H-score 155.3±54.19 vs 137.5±60.72, *P* value 0.5444; Supplementary Figure 8). Collectively, these data suggest that the deletion is unlikely to affect local chromatin interaction patterns and the expression of neighboring genes within affected tissues.

Interestingly, *SLC30A9* mRNA was expressed at similar levels in cells from affected patients compared with healthy controls suggesting, together with RT-PCR data on peripheral blood samples (Supplementary Figure 3B), that mutant mRNA may evade nonsense-mediated decay (NMD). This was indeed confirmed by the absence of mutant mRNA accumulation in patient-derived lymphoblastoid cells following treatment with cycloheximide, a potent NMD inhibitor (Figure 4B)^26^. Upon expressing the mutant transcript with differential terminal tags in HEK293T cells, the predicted short protein p.(Glu37Alafs*11) could not be detected (data not shown) – likely due to its instability – yet blotting for the C-terminal FLAG tag allowed us to identify two truncated proteins of approximately 30-40kDa (Figure 4C). These products could also be detected with an SLC30A9-specific antibody (Figure 4D), suggesting that they represent in-frame truncated SLC30A9 proteins. The size of these products was consistent with translation reinitiation from in-frame AUG codons located downstream of the deletion in exons 8 (Met241) and exon 9 (Met266; Figure 4A). To confirm this, introduction of a stop codon at position 258 resulted in loss of the largest protein product (Δ240 SLC30A9), while introducing a stop codon at position 269 caused loss of both proteins (Δ240 and Δ265 SLC30A9) (Figure 4C). The localization of these N-terminally truncated protein products was investigated using immunofluorescence. Consistent with published data^27,28^, wild-type (WT) SLC30A9 was found to colocalize with the mitochondrial marker TOMM20. However, mutant SLC30A9 showed loss of colocalization with TOMM20 (Supplementary Figure 9), in keeping with the absence of the N-terminal mitochondrial presequence^29^ in the truncated forms. Mutant SLC30A9 showed discrete peri-nuclear dot-like expression which co-localized with calreticulin (Figure 4E), suggesting aggregation of the mutant proteins within the ER. Furthermore, mutant SLC30A9 showed decreased turnover compared to WT SLC30A9 in cycloheximide chase experiments (Figure 4F).

**Figure 4.**
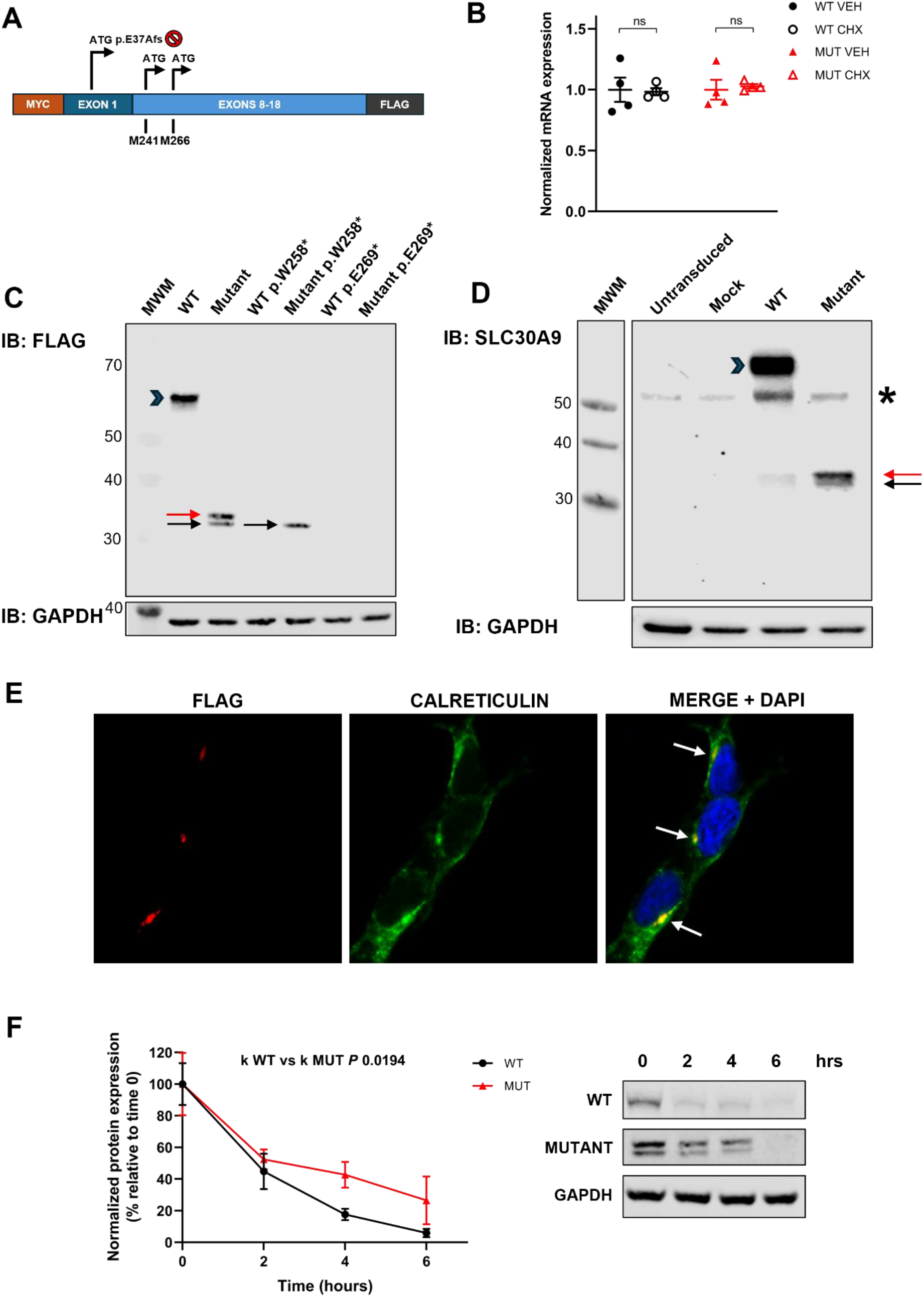
The *SLC30A9* intragenic deletion results in translation reinitiation and production of truncated proteins localizing to the endoplasmic reticulum. A. Schematic of the mutant *SLC30A9* construct. The stop codon in exon 8 is shown with a red asterisk. In-frame AUG codons downstream of the deletion (Met241 and Met266) are highlighted. B. Patient-derived EBV-transformed lymphoblastoid cells were treated with vehicle (VEH) or cycloheximide (100mcg/ml; CHX) for six hours. The expression of wild-type (WT) and mutant (MUT) mRNA was quantified using RT-qPCR. There was no evidence of accumulation of the mutant transcript following CHX treatment, suggesting evasion from nonsense-mediated decay (multiple t tests; data shown as individual values, mean, and SEM, n = 4). C. Transient expression of WT FLAG-tagged *SLC30A9* in HEK293T cells results in a full-length protein of approximately 60kDa (blue arrowhead), as shown by immunoblotting (IB) for FLAG. Expression of mutant *SLC30A9* results in two N-terminally truncated products of approximately 30-40kDa, consistent with Δ240 (red arrow) and Δ265 SLC30A9 (black arrow). Introducing a stop codon at position 258 leads to loss of Δ240, while a stop codon at position 269 leads to loss of both products, in keeping with translation reinitiation at codons 241 and 266. MWM = molecular weight marker. D. The expression of SLC30A9 was investigated by immunoblotting (IB) in untransduced cells and TT cells infected with mock, WT, and mutant SLC30A9 lentiviruses. The full-length protein is indicated by a blue arrowhead, while the truncated proteins are highlighted by red and black arrows. The asterisk highlights endogenous SLC30A9. Low-level expression of the truncated products was also noted when overexpressing WT SLC30A9, although these were not detected in endogenous conditions. E. Mutant SLC30A9 showed dot-like perinuclear expression which colocalized with the endoplasmic reticulum marker calreticulin (HEK293T cells). F. HEK293T cells stably expressing WT and mutant SLC30A9 were treated with cycloheximide (20μg/ml) for the indicated time points. The top panel shows representative Western blot images. The lower graph shows the degradation curves obtained after plotting normalized protein levels. The degradation speed (k) of the mutant proteins was significantly reduced compared to WT SLC30A9 (extra sum-of-squares *F* test; n=4).

We next introduced the *SLC30A9* deletion into TT cells using CRISPR and analyzed the transcriptome of mutant cells with RNA-Seq. In comparison to a control clone, mutant cells showed significant enrichment of genes involved in ‘ECM-receptor interaction’, ‘phagosome’, ‘protein processing in the endoplasmic reticulum’, and ‘lysosome’ KEGG pathways (Figure 5A). There was significant upregulation of genes involved in the unfolded protein response (UPR), including ER chaperones *HSPA5* and *HSP90B1*, as well as the ER-associated degradation (ERAD) gene *EDEM2,* lysosomal proteases including *CTSA*, *CTSD*, and *CTSL*, and the autophagy-inducing *MCOLN1* gene (Figure 5B and Supplementary Table 8). There was no difference in the expression of *RET* (data not shown). The PI3K-Akt signaling pathway was also significantly upregulated in mutant cells, in keeping with the known cross-talk between the UPR and the PI3K-Akt pathway^30,31^. We confirmed upregulation of GRP78 (protein product of *HSPA5*) at the protein level (Figure 5C). Given published evidence linking the *MET* oncogene to the pro-tumorigenic effects of the UPR^32^, we assessed the protein expression of MET and found it to be significantly overexpressed in mutant cells (Figure 5C). Lastly, mutant TT cells were found to have significantly increased cell proliferation (Figure 5D), clonogenic potential (Figure 5E), and cell migration (Figure 5F), strongly supporting the pathogenic role of the *SLC30A9* deletion.

**Figure 5.**
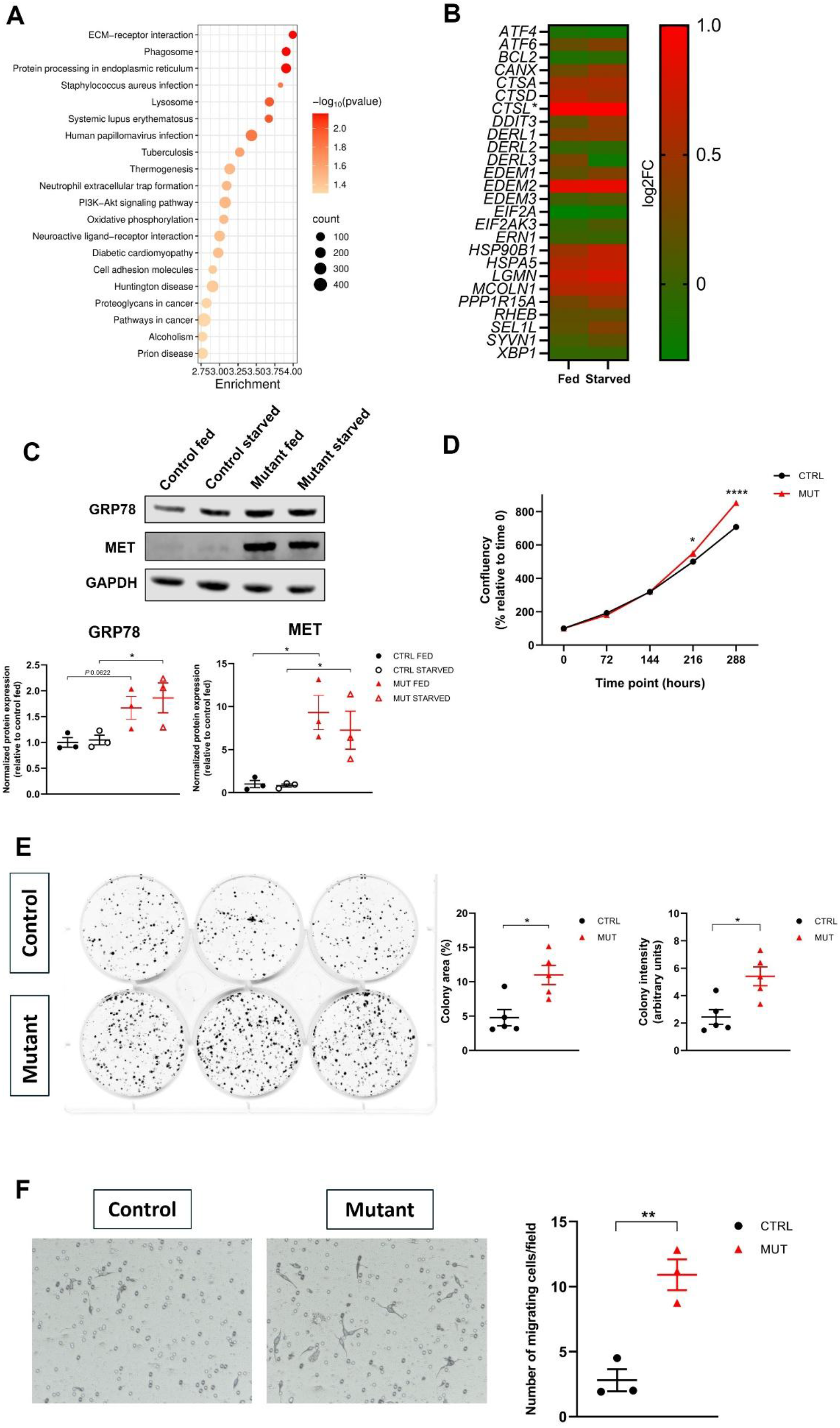
The *SLC30A9* intragenic deletion results in activation of the unfolded protein response pathway and has oncogenic effects in MTC cells. A. Dot plot graph of RNA-Seq data in CRISPR-edited TT cells following acute serum starvation (3 hours) showing, among others, positive enrichment of the unfolded protein response and the PI3K-Akt signaling pathways in mutant cells. B. Heatmap analysis of genes involved in the unfolded protein response, ER-associated degradation, and lysosomal degradation pathways in CRISPR-edited TT cells (mutant vs control) in fed and starved conditions. Log2FC, log2 fold change. \**CTSL* log2FC was higher than the scale provided (see Supplementary Table 8). C. Upregulation of the endoplasmic reticulum chaperone GRP78, one of the main regulators of the unfolded protein response, was confirmed at the protein level. Marked overexpression of the MET oncogene was also detected in both fed and starved conditions (Two-Way ANOVA with Tukey’s multiple comparison test; data shown as individual values, mean, and SEM, n = 3). D. CRISPR-edited mutant TT cells showed significantly increased cell proliferation compared with control cells as assessed by live-imaging confluency (Two-Way ANOVA with Sidak’s multiple comparisons test; data shown as mean with SEM, n=3). E. The mutation significantly increased clonogenic capacity in TT cells as shown by colony forming assays. The left panel shows representative images. The graphs on the right show colony area and intensity (Mann-Whitney test; data shown as individual values, mean, and SEM, n=5). E. Mutant cells showed increased migration as compared to control. Representative images are shown (10x magnification) (unpaired t test; data shown as individual values, mean, and SEM, n=3).

## Discussion

CNVs account for approximately 10% of pathogenic mutations in cancer susceptibility syndromes^33,34^. While most of these variants are thought to result in loss-of-function, there are limited data on alternative mechanisms linking structural genomic variants to tumor development. Here, we describe a unique *SLC30A9* intragenic deletion as a novel cause of fMTC in two related families with 21 affected individuals over four different generations. This represents the largest described kindred with fMTC in which a pathogenic *RET* mutation could not be identified as the cause of the disease.

Biallelic loss-of-function small variants in *SLC30A9* have been linked to Birk-Landau-Perez syndrome^17–22^, a condition characterized by a predominantly neurological phenotype, suggesting that loss of SLC30A9 function primarily affects the central nervous system. Importantly, there is no evidence of thyroid cancer in subjects with Birk-Landau-Perez syndrome or in the heterozygous carriers within these families, further confirming that the fMTC phenotype is not dependent on SLC30A9 loss-of-function. We undertook comprehensive analyses to exclude a role for genes other than *SLC30A9* from the linkage region at chromosome 4p13 and to address the possibility of regulatory effects of the deletion on the expression of closely located, preserved genes in normal and MTC tumor cells from affected family members and controls. No other pathogenic variants were identified in genes within the linkage region, and chromatin conformation capture experiments showed no evidence of effects on the regulation of neighboring genes.

Our functional studies provide evidence that the intragenic *SLC30A9* deletion does not act through a classic loss-of-function mechanism. In fact, knockdown of *SLC30A9* in an MTC cell line had no effect on cell viability or major oncogenic signaling pathways. Instead, the mutation resulted in the production of N-terminally truncated proteins via NMD evasion and translation reinitiation. These truncated proteins localized within the ER, and introducing the deletion in a human MTC cell line was associated with increased cell proliferation, colony forming, and migration capacities, alongside positive enrichment of the UPR and PI3K-Akt pathways. NMD is a conserved mRNA surveillance system responsible for degrading transcripts with premature stop codons to prevent the production of truncated proteins^35,36^. However, certain stop codons fail to induce NMD, resulting in translation from nearby AUG codons. It has been shown that premature stop codons residing in the last exon or at least 50 nucleotides (nt) 5’ to the last exon-exon junction evade NMD^37,38^. Additionally, premature stop codons within the first 150nt or those located in exons longer than 407nt also tend to evade NMD^38^, highlighting the critical role of the stop codon position in determining NMD activation. In our case, NMD evasion is likely to occur as a result of the ‘start-proximal rule’, as the premature stop codon resulting from the exon 2-7 deletion falls within the first 150nt of the mutant transcript, therefore allowing mRNA translation from downstream in-frame AUG codons. Collectively, our findings implicate NMD evasion and translation reinitiation as the causative mechanisms linking the *SLC30A9* deletion with fMTC. Interestingly, translation reinitiation has been confirmed experimentally for several pathogenic nonsense variants involving the *TP53* gene, suggesting it may represent an under-detected phenomenon^39^. There are reported examples of germline variants in other tumor suppressor genes which disrupt the canonical start codon and result in translation initiating from alternative AUG codons^40^, with loss of the normal protein function. However, to the best of our knowledge, this represents the first report of translation reinitiation and activation of the UPR as the mechanisms involved in a hereditary cancer susceptibility syndrome.

The UPR is a collection of adaptive signaling pathways aimed at maintaining a functioning ER protein folding environment^41^. Several lines of evidence support a role for the UPR in all stages of cancer development, including transformation, progression, and metastasis^42–45^. UPR activation in cancer cells can be attributed to oncogenes or loss of tumor-suppressor genes^46–49^, as well as increased protein synthesis owing to the high metabolic demands linked with oncogenic transformation. Additionally, certain oncogenic mutations result in the retention of misfolded proteins within the ER with intrinsic activation of the UPR programme^50^. Misfolded proteins bind the chaperone GRP78, and the consequent reduction in free GRP78 activates the PERK-ATF4, IRE1α-XBP1, and the ATF6 pathways. The induction of the UPR is aimed at reducing protein synthesis, increasing the expression of chaperones facilitating protein folding, as well as promoting protein degradation pathways including ERAD and autophagy. The tumorigenic effects of the UPR are likely multifactorial. GRP78 itself has been shown to localize to the cell membrane in several cancer cells and act as a receptor for pro-proliferative pathways^51–53^. ATF6 has also been shown to activate the PI3K-Akt pathway, promoting cell proliferation^54,55^. Additionally, MET oncogene overexpression has been linked with the integrated stress response, and is likely to contribute to the growth promoting effects of the UPR in cancer cells^32^. In case of severe and prolonged activation of the UPR, pro-apoptotic signals are activated predominantly through the PERK-ATF4 and IRE1α-XBP1 pathways, highlighting the dual role of the UPR as both an adaptive and pro-apoptotic process.

Our findings provide evidence that the *SLC30A9* intragenic deletion results in an adaptive UPR programme in MTC cells. This is confirmed by the positive pathway enrichment and upregulation of several key ER chaperones (including *HSPA5*) and the ERAD-associated *EDEM2* gene, whose overexpression has been linked with unfavourable prognosis in malignant gliomas^56^. The concomitant upregulation of lysosomal proteases and the autophagy-inducing *MCOLN1* gene^57^ further suggests increased capacity for protein degradation, while the reduced turnover of the truncated proteins reflects the paradoxical negative effects of protein synthesis inhibition on the degradation of unfolded proteins^58,59^. Positive enrichment of the PI3K-Akt pathway and significant upregulation of MET in mutant cells are also likely to underlie the pro-proliferative effects of the deletion in MTC cells. The significant overexpression of MET is also relevant in view of previous findings linking a germline *MET* variant with *RET* mutation-negative fMTC in one kindred^14^, implicating this oncogene in the pathogenesis of the disease. *ATF4* and *XBP1* were not significantly induced in either fed or starved conditions in mutant cells, further highlighting an adaptive response rather than strongly sustained ER stress. This is further confirmed by the lack of significant upregulation of *DDIT3*, a key effector of ER stress-induced apoptosis^60^, and no changes in *BCL2* expression.

The *SLC30A9* gene encodes a 568 amino acid protein which is expressed in a variety of human tissues and cancer cell lines^61^. SLC30A9 (also known as ZnT9) belongs to the SLC30 (or ZnT) family of zinc transporters and has been shown to maintain mitochondrial zinc homeostasis^27–29,62,63^. It is unique among other members of this family of zinc transporters, as it presents several features that also support its ability to translocate to the nucleus and act as a transcriptional co-regulator for multiple pathways^61,64–67^. Recent studies highlighted an oncogenic role of SLC30A9: overexpression was associated with worse outcomes in diffuse large B cell lymphoma^68^ and increased cell proliferation and migration in cervical squamous cell carcinoma^69^. The relevance of these observations to the fMTC phenotype is uncertain. Furthermore, the reasons underlying the selectivity of the phenotype despite the relatively broad expression pattern of *SLC30A9* are unclear. It has been noted that cells with high secretory capacity, such as neuroendocrine cells, are particularly reliant on the adaptive UPR^70–72^, providing a possible functional link between the *SLC30A9* deletion and MTC.

The age of disease onset in the two families aligns with that described for families with ‘moderate risk’ *RET* mutations^73^, and suggests that close monitoring of calcitonin levels should be undertaken in *SLC30A9* deletion carriers starting around the age of 5 years. Notwithstanding the obligate carriers who could not be screened clinically, disease penetrance was virtually complete, and it is therefore expected that all carriers will require a thyroidectomy. While exact timing of surgery remains to be established, clinical experience in these families suggests that it would be sensible for carriers to undergo thyroidectomy at the time calcitonin levels become elevated, which should predate the occurrence of clinically significant thyroid cancer.

No *SLC30A9* CNVs or small variants were identified in seven further families with fMTC or fMTC-like phenotype and in a cohort of patients with sporadic MTC, suggesting that the *SLC30A9* intragenic deletion is likely to represent a private mutation, much akin to previously reported fMTC/fMTC-like cases harboring mutations in the *ESR2*^13^ and *MET*^14^ genes. These findings broaden our understanding of the genetic basis of MTC, and confirm that multiple genes contribute to disease susceptibility beyond *RET*.

Several limitations should be considered in this study. The *SLC30A9* deletion was identified in only two related families, limiting assessment of the phenotypic spectrum and recommendations on management. Furthermore, functional studies were performed in the TT MTC cell line and therefore may not fully recapitulate the biology of MTC *in vivo*. Lastly, although effects on mitochondrial zinc homeostasis are unlikely given the heterozygous state of the mutation and lack of co-localization of the mutant proteins with mitochondrial markers, further experiments looking at potential effects on zinc metabolism will be needed to fully dissect the role of SLC30A9 in MTC.

## Methods

### Patients

We recruited nine families with *RET* mutation-negative fMTC (n=7) or fMTC-like phenotype (n=2; i.e. one case of MTC and at least one case of pathologically-proven C cell hyperplasia in a first-degree relative), and 108 patients with sporadic MTC without germline *RET* mutations through the multicenter “Genetics of endocrine tumours” study approved by the National Research Ethics Service Committee East of England – Cambridge East (REC reference 06/Q0104/133). All patients provided a signed consent form. All patients provided a sample for DNA extraction (either saliva or an EDTA blood tube). Clinical data were provided by the referring physicians. When available, formalin-fixed paraffin-embedded tumor samples were also obtained. Details on DNA extraction and *RET* testing procedures are provided in Supplementary Methods.

### Linkage analysis

SNP genotyping was performed in 40 individuals from the two index families (35 from family 1 and five from family 2) using the Illumina CytoSNP12-BeadChip (Illumina, San Diego, CA, USA). Genotyping quality control and assessment of relatedness were performed using PLINK (https://www.cog-genomics.org/plink/) and a PI_HAT threshold of 0.05 was applied to establish relatedness, as previously suggested^74^. Pedigree errors or cryptic relatedness were assessed using Prest-plus (https://utstat.toronto.edu/sun/Software/Prest/). The LINKDATAGEN tool was used to generate linkage style files (https://bioinf.wehi.edu.au/software/linkdatagen/), and MORGAN (https://faculty.washington.edu/eathomp/Genepi/MORGAN/Morgan.shtml) was employed for linkage analysis. A LOD score ≥3 was considered indicative of genetic linkage, as previously suggested^75^. Haplotypes were reconstructed using SHAPEIT (https://mathgen.stats.ox.ac.uk/genetics_software/shapeit/shapeit.html) and visualized with HaploPainter (https://haplopainter.sourceforge.net/).

### Exome and genome sequencing

Exome sequencing was performed using the SureSelect Human All Exon platform (V4+UTRs) (Agilent, Santa Clara, CA, USA) in four affected subjects from the index families with sequencing on a HiSeq2500 system (Illumina). Mean coverage was 183.4 (family 1: IV/1), 174.9 (family 1: IV/6), 182.7 (family 1: V/7), and 143.1 (family 2: III/2). Exome sequencing data were analyzed using the Varsome platform (https://landing.varsome.com/varsome-premium). Genome sequencing was performed under standard conditions in two affected subjects from the index families on a HiSeq2500 system (Illumina). Mean coverage was 35 (family 1: IV/16) and 44.1 (family 2: IV/1). Samtools^76^, Varscan^77^, and GATK^78^ were used for variant calling. NovoBreak^79^, Delly^80^, and Manta^81^ were used to detect structural variants. The effect of intronic variants on splicing was investigated using Human Splicing Finder (https://bio.tools/human_splicing_finder) and SpliceAI (https://spliceailookup.broadinstitute.org/).

### High-definition array comparative genomic hybridization

A custom HD-aCGH targeting chromosome 4 between position 15 and 70Mb (GRCh37/hg19) was designed (http://earray.chem.agilent.com/suredesign). The array comprised 166,417 total probes with a median probe spacing of 221bp (4 x 180K format). Protocols for labeling DNA samples, hybridization of the arrays, and data analysis were as previously described^82^.

### Multiplex ligation-dependent probe amplification

A custom MLPA probe mix was designed by MRC Holland (Amsterdam, the Netherlands) to assess *SLC30A9* copy number. Details on probe composition and analytical procedures are provided in Supplementary Methods.

### Targeted sequencing

*SLC30A9* (NM_006345.4) and *RET* (NM_020975.6) were sequenced in probands from the *RET* mutation-negative MTC families and all patients with *RET* mutation-negative sporadic MTC using the Fluidigm Access Array (Fluidigm, South San Francisco, CA, USA). Further details are provided in Supplementary Methods.

### PCR, RT-PCR, and Sanger sequencing

PCR and Sanger sequencing were employed to define the breakpoints of the *SLC30A9* intragenic deletion. Co-segregation of the deletion with the disease trait was assessed by duplex PCR in available members of families 1 and 2. The effect of the deletion on *SLC30A9* splicing was assessed by RT-PCR of RNA extracted from peripheral blood leukocytes of five deletion carriers. Somatic *RET* and *RAS* hotspot mutations were assessed by PCR and Sanger sequencing of gDNA from available formalin-fixed paraffin-embedded tumor samples. Primer sequences are available upon request. Further details on nucleic acid extraction are provided in Supplementary Methods.

### OncoScan assay

The OncoScan assay (Affymetrix, Santa Clara, CA, USA) was employed to detect copy number changes in DNA isolated from formalin-fixed paraffin-embedded tumor samples from family 1. The assay was run on a GeneChip scanner 3000 (AppliedBiosystems, Foster City, CA, USA) according to the manufacturer’s instructions. Data analysis was performed using the Nexus Copy Number software (v10, BioDiscovery, El Segundo, CA, USA).

### Cell culture

HEK293T and TT cells were acquired from ECACC (Salisbury, UK) and grown in their respective recommended media supplemented with 10% fetal bovine serum (Thermo Fisher Scientific, Waltham, MA, USA) and penicillin-streptomycin (Thermo Fisher Scientific). EBV-transformed lymphoblastoid cell lines were established from peripheral blood lymphocytes from three patients from family 2 (III/2, IV/1, and IV/4) and one healthy control by ECACC. Skin fibroblast primary cultures were established from three patients from family 2 (III/2, IV/1, and IV/4) via skin punch biopsy and cell disassociation. All cell lines were cultured in a humidified incubator containing 5% CO_2_ at 37°C and were routinely tested for Mycoplasma with the MycoAlert® Mycoplasma Detection Kit (Lonza, Basel, Switzerland).

### RNA-Seq

RNA was extracted from lymphoblastoid cell lines, skin fibroblasts, and CRISPR-modified TT cells from six different passages using the RNeasy Micro Kit (Qiagen, Hilden, Germany) and submitted for RNA-Seq following quality control analysis with an Agilent 2100 Bioanalyzer (Agilent). FastQC version 0.11.9 was used for quality control of the reads data. HISAT2 version 2.2.1^83^ was used for read alignment. Sam Tools version 1.10 was employed to convert sam files to sorted and indexed bam files and Picard Metrics version 2.26.6 for further quality control. Quantification of the RNA-sequencing data as counts per gene was completed using subread_featureCounts version 2.01^84^. R studio was used for differential gene expression analysis using DESeq2^85^. An adjusted *P* value <0.05 and log2 fold change of +/-0.5 were considered significant for differential gene expression analysis. GAGE was employed for pathway analysis in CRISPR-edited cells using a false discovery rate (FDR) cut-off of <0.05^86^.

### Chromosome conformation capture experiments

Nuclei isolation and Hi-C library construction in TT cells, using the DpnII restriction enzyme, followed a previously established protocol^23^ with minor modifications detailed here^87^. The final Hi-C libraries were sequenced using DNBseq technology, producing 100bp paired-end reads, with each sample yielding approximately 400 million raw sequencing read pairs. Hi-C data analysis was performed as previously described with minor modifications^88^. UMI-4C library preparation was carried out as previously described^25^, with modifications in the 3C library preparation and sequencing library construction, as detailed previously^87^. Experiments were performed in singleton using skin-derived fibroblasts from one affected subject and a healthy control sample. Further details are provided in Supplementary Methods.

### Lentiviral vectors and plasmids

Codon-optimized, MYC-(N-terminus), and FLAG-tagged (C-terminus) WT and mutant (exons 2-7 deletion) *SLC30A9* subcloned in the pcDNA3.1+/C-DYK vector were obtained from GenScript Biotech (Piscataway, NJ, USA). With the aim to assess the sites of translation reinitiation, stop codons were introduced at codons 258 and 269 using the QuikChange II XL Site-Directed Mutagenesis Kit (Agilent). Primer sequences are available upon request.

The GIPZ Transduction Starter Kit (Horizon Discovery, Cambridge, UK) was used to generate small hairpin RNAs (shRNAs) targeting *SLC30A9*. Sequences are available upon request. WT and mutant *SLC30A9* were subcloned into the pLOC lentiviral vector (Horizon Discovery, Cambridge, UK) and lentiviruses for stable expression were produced in HEK293T cells under standard conditions. HEK293T and TT cells were transduced using hexadimethrine bromide (Sigma-Aldrich, Burlington, MA, USA) with subsequent antibiotic selection. A mock control lentivirus (empty vector) was also included.

### CRISPR editing

The *SLC30A9* deletion was introduced into TT cells using the lentiCRIPSR v2 vector (kindly supplied by Dr Chris Smith, QMUL) and two subsequent transductions with gRNAs targeting intron 1 and intron 7, respectively. gRNA sequences are available upon request. Single cell clones were selected following CellenONE sorting (Cellenion, Lyon, France) and successful editing was confirmed at both gDNA and cDNA level. A control clone was established from cells transduced with a gRNA targeting intron 1 only.

### Western blotting

Western blotting was performed under standard conditions and with the following primary antibodies: anti-SLC30A9 (Santa Cruz Biotechnology, Dallas, TX, USA; sc-271956), anti-MYC (Santa Cruz Biotechnology; sc-40), anti-FLAG (Sigma-Aldrich; F1804), anti-pERK (Sigma-Aldrich; M8159), anti-ERK (Sigma-Aldrich; M5670), anti-pAKT(S473) (Cell Signaling Technology, Danvers, MA, USA; 4060), anti-AKT (Cell Signaling Technology; 4691), anti-GRP78 (Abcam, Cambridge, UK; ab21685), anti-MET (Cell Signaling Technology; 3127), and anti-GAPDH (Abcam; ab181602).

### Cycloheximide experiments

To assess whether mutant *SLC30A9* mRNA evades NMD, EBV-transformed lymphoblastoid cells from one deletion carrier and one healthy control were treated with cycloheximide (100μg/ml; Sigma-Aldrich) for six hours. WT and mutant mRNA were then quantified via RT-qPCR using the Brilliant III Ultra-Fast SYBR green master mix (Agilent) on an Agilent AriaMx machine (Agilent). Primer sequences are available upon request.

The stability of WT and mutant SLC30A9 was assessed in stably expressing HEK293T cells following treatment with cycloheximide (20μg/ml; Sigma-Aldrich) for the indicated time points. Results were analyzed using a one-phase decay equation. The degradation constant (K) was compared between WT and mutant proteins using the extra sum-of-squares *F* test, as previously described^89^.

### Cell viability, proliferation, colony forming, and cell migration assays

Alamar blue (Thermo Fisher Scientific) fluorescent dye was used to measure viability in TT cells following knockdown of *SLC30A9*. For CRISPR-modified TT cells, cell proliferation was assessed by means of live-cell microscopy using an IncuCyte S3 device (Sartorius, Göttingen, Germany). Clonogenic capacity was assessed with colony forming assays. Cell migration was assessed using BioCoat Control Inserts (Corning Inc., Corning, NY, USA). Further details are provided in Supplementary Methods.

### Immunofluorescence

Stably expressing HEK293T cells were seeded in an 8-well Lab-Tek II Chamber Slide (Sigma-Aldrich). Following fixation with methanol, immunofluorescence was carried out under standard conditions and the following primary antibodies: anti-FLAG (Sigma-Aldrich; F1804), anti-TOMM20 (Proteintech, Rosemont, IL, USA; 11802-1-AP), and anti-calreticulin (Abcam; ab92516). Images were obtained with an LSM880 confocal microscope (Zeiss, Oberkochen, Germany). Colocalization between SLC30A9 and TOMM20 was assessed using the ImageJ software and the Cololization_Finder plugin (http://questpharma.u-strasbg.fr/html/colocalization-finder.html).

### Immunohistochemistry

Immunohistochemistry for SLC30A9, SHISA3, calcitonin, and Ki-67 was performed on available formalin-fixed paraffin-embedded tumor samples. The following primary antibodies were employed: anti-SLC30A9 (Santa Cruz Biotechnology; sc-271956), anti-SHISA3 (Thermo Fisher Scientific; PA5-34527), anti-calcitonin (Novus Biologicals, Littleton, CO, USA; NBP2-21752), and anti-Ki-67 (Abcam; ab16667). Immunostainings were performed on 4µm tissue sections using the Ventana Discovery DAB Map System (Ventana, Oro Valley, AZ, USA). Reactions where the primary antibodies were omitted were included as negative controls. The Congo Red stain was performed at the Pathology Core Facility of the Blizard Institute (Queen Mary University of London) under standard conditions. The expression of SLC30A9 and SHISA3 was assessed semi-quantitatively taking into account both staining intensity (range 0-3) and the percentage of positive cells (H-score, range 0-300), as previously described^90^. The Ki-67 proliferation index was based on the evaluation of at least 500 tumor cells in areas of highest nuclear labeling (‘hotspots’). Scoring of immunohistochemistry was performed by two pathologists (D.I. and L.O.).

### Statistics

Clinical data were normally distributed and are presented as mean ± SD. Experimental data are presented as mean ± SD or graphically as mean with SEM. Normal distribution was assessed using the Shapiro-Wilk test. Data were compared through parametric or non-parametric tests as appropriate using the Prism v8 software (GraphPad Software). Significance was assumed for *P* values <0.05.

## Data Availability

All data produced in the present study are available upon reasonable request to the authors.

## Ethics

All patients provided signed consent forms, and the study was approved by the National Research Ethics Service Committee East of England – Cambridge East (REC reference 06/Q0104/133).

## Data and materials availability

Cell lines used in this study are available upon request.

## Author contributions

D.I. and M.K. conceived the study and D.I., F.B., C.M.B.C., M.F., R. K., J.R.L., J.R.S., L.D.M., and M.K. designed research. D.I., F.B., and M.K. acquired funding for the project. D.I. wrote the manuscript with input from F.B. and M.K. D.I., F.B., and O.S. performed laboratory-based experiments. M.D., A.C., M.M., H.S., Me.T., and R.K. performed linkage analyses and analyzed data. D.I., K.S., S.A., S.B., C.P.C., C.L.H., K.O.K., L.E.T., and M.W. analyzed next-generation sequencing data. K.C. contributed to prospective patient testing. J.M.F., M.G., C.M.G., J.P., and C.M.B.C. designed the HD-aCGH, performed testing, and analyzed data. S.L. and S.Sa. designed the MLPA assay, performed testing, and analyzed data. S.Sc. and S.W.W. performed *in silico* analyses on chromatin conformation data. M.F. performed chromosome conformation capture experiments and analyzed data. D.I., M.A., A.B., C.B., F.C., S.C., M.R.D., P.D., W.D.F., B.J., J.K., T.K., F.L., K.M., Re.M., S.M., J.S.M., Ra.M., P.J.M., M.O-W., M.O., A.P., S.P., A.R., P.R., D.M.S-C., Mi.T., J.R.S., L.D.M., and M.K. recruited patients and contributed to sample and data collection. All authors reviewed the manuscript.

## Acknowledgments

We acknowledge all participating patients for their contributions to this study. Dr Gabor Czibik (Queen Mary University of London) is gratefully acknowledged for assistance with experimental plan design. We acknowledge the Department of Endocrinology at Aalborg University Hospital (Aalborg, Denmark) for patient consenting and recruitment, and the late Professor Charis Eng for contributing to the recruitment of sporadic MTC patients.

