## Supplementary material for "Familial medullary thyroid carcinoma secondary to an *SLC30A9* intragenic deletion and translation reinitiation"

| Patient ID | Gender | Age at diagnosis (range) | Calcitonin at diagnosis (normal range) | Surgery | TNM staging (adapted to 9th edition UICC) | Treatment outcome (remission, persistent, recurrent disease) | Calcitonin doubling time (if persistent/recurrent disease) | Notes |
|---|---|---|---|---|---|---|---|---|
| F1 II/1 | M | NA | NA | NA | NA | NA | | Obligate carrier, no clinical data available |
| F1 II/3 | F | 46-50 | NA | No | | | | Died of thyroid cancer in her 40s |
| F1 III/1 | F | 41-45 | NA | NA | NA | NA | | Died of MTC in her 40s |
| F1 III/3 | F | 36-40 | NA | Yes | pT3(m) pN0 | Remission | | |
| F1 III/6 | M | | | | | | | Obligate carrier; refused clinical screening |
| F1 III/9 | M | | | | | | | Obligate carrier; died of esophageal cancer |
| F1 IV/1 | F | 21-25 | NA | Yes | pT1a(m) pN0 | Remission | | |
| F1 IV/2 | F | 16-20 | 179pg/ml (<14) | Yes | pT1?(m) pN1b | Persistent disease | >24 months | Had repeat lymphadenectomy for metastatic MTC in her 20s |
| F1 IV/6 | F | 16-20 | NA | Yes | pT? pN0 | Remission | | |
| F1 IV/9 | F | 31-35 | 325pg/ml (<14) | Yes | pT1b(m) pNx | Remission | | |
| F1 IV/14 | M | 21-25 | 168pg/ml | Yes | pT1a(m) pN1a | Remission | | |
| F1 IV/16 | M | 21-25 | 200pg/ml | Yes | pT1a(m) pN0 | Remission | | |
| F1 IV/19 | M | 16-20 | NA | Yes | NA | Remission | | |
| F1 V/4 | M | 16-20 | 740pg/ml (<18.2) | Yes | pT2(m) pN0 | Remission | | |
| F1 V/5 | M | 21-25 | 32.8pg/ml (<18.2) | Yes | pT1a(m) pN1a | Remission | | |
| F1 V/7 | F | 11-15 | 64pg/ml (<11.5) | Yes | pT1a(m) pN0 | Remission | | |
| F2 I/1 | M | NA | NA | Yes | NA | NA | | Was known to have had neck surgery at young age |
| F2 II/2 | F | 6-10 | NA | Yes | NA | NA | | Died aged 6-10, history of neck surgery |
| F2 II/4 | F | 31-35 | NA | Yes | NA | Persistent disease | NA | Died of MTC in her 30s |
| F2 III/2 | F | 31-35 | NA | Yes | pT? pN1b | Recurrent disease | >24 months | Had repeat lymphadenectomy x2 for metastatic MTC; concurrent primary hyperparathyroidism (parathyroid adenoma) |

| | | | | | | | |
|---|---|---|---|---|---|---|---|
| F2 IV/1 | F | 16-20 | NA | Yes | NA | Remission | |
| F2 IV/4 | M | 6-10 | 22pg/ml (<8.4) | Yes | pT1a(m) pNx | Recurrent disease | >24 months |
| **Affected subjects identified prospectively following genetic testing** | | | | | | | |
| F1 III/7 | F | 61-65 | 625.8pg/ml (<9.8) | Yes | pT1b(m) pN1b | Remission | |
| F1 V/1 | F | 16-20 | 14.3pg/ml (<11.5) | Yes | pT1a(m) pNx | Remission | |

**Supplementary Table 1. Clinical features of patients diagnosed with MTC in the two index families.** F1, family 1; F2, family 2; NA, not available; UICC, Union for International Cancer Control.

| Gene name | Start position (hg19) | End position (hg19) | RefSeq match | Description |
|---|---|---|---|---|
| *TMEM33* | 41,937,446 | 41,962,820 | NM_018126.3 | Transmembrane Protein 33 |
| *DCAF4L1* | 41,983,773 | 41,988,482 | NM_001029955.4 | DDB1 And CUL4 Associated Factor 4-like 1 |
| *SLC30A9* | 41,992,547 | 42,092,478 | NM_006345.4 | Solute Carrier Family 30 Member 9 |
| *BEND4* | 42,112,870 | 42,154,672 | NM_207406.4 | BEN Domain Containing 4 |
| *SHISA3* | 42,399,505 | 42,404,504 | NM_001080505.3 | Shisa Family Member 3 |
| *ATP8A1* | 42,410,390 | 42,659,122 | NM_006095.2 | ATPase Phospholipid Transporting 8A1 |
| *GRXCR1* | 42,894,730 | 43,032,675 | NM_001080476.3 | Glutaredoxin And Cysteine Rich Domain Containing 1 |
| *KCTD8* | 44,175,920 | 44,450,826 | NM_198353.3 | Potassium Channel Tetramerization Domain Containing 8 |
| *YIPF7* | 44,624,105 | 44,653,642 | NM_182592.3 | Yip1 Domain Family Member 7 |
| *GUF1* | 44,680,437 | 44,702,945 | NM_021927.3 | GTP Binding Elongation Factor GUF1 |
| *GNPDA2* | 44,703,812 | 44,728,651 | NM_138335.3 | Glucosamine-6-Phosphate Deaminase 2 |
| *GABRG1* | 46,037,786 | 46,126,071 | NM_173536.4 | Gamma-Aminobutyric Acid Type A Receptor Subunit Gamma1 |
| *GABRA2* | 46,245,565 | 46,392,317 | NM_000807.4 | Gamma-Aminobutyric Acid Type A Receptor Subunit Alpha2 |
| *COX7B2* | 46,736,844 | 46,911,262 | NM_130902.3 | Cytochrome C Oxidase Subunit 7B2 |

**Supplementary Table 2. Details of the 11 protein-coding genes present within the shared haplotypes in the index families.** Genes in the gap between the shared haplotypes are highlighted in light gray.

| Variant (hg19) | Gene | Transcript | Coding impact | Allelic balance F1 V/7 | Allelic balance F1 IV/6 | Allelic balance F1 IV/1 | Allelic balance F2 III/2 | ACMG interpretation | Varsome *in silico* predictions (benign vs pathogenic) | Coverage F1 V/7 | Coverage F1 IV/6 | Coverage F1 IV/1 | Coverage F2 III/2 |
|---|---|---|---|---|---|---|---|---|---|---|---|---|---|
| chr11:63536006 A⇒G | *ZFTA* | NM_001144936.1 c.89T>C p.L30P | missense | 0.390625 | 0.47619 | 0.321429 | 0.423077 | Uncertain significance | Benign (7 vs 0) | 64 | 21 | 56 | 26 |
| chr11:118307393 T⇒C | *KMT2A* | NM_005933.3 c.166T>C p.S56P | missense | 0.225806 | 0.45 | 0.3125 | 0.391304 | Uncertain significance | Benign (9 vs 3) | 31 | 20 | 16 | 23 |
| chr19:48994708 T⇒G | *LMTK3* | NM_001080434.1 c.4268A>C p.H1423P | missense | 0.333333 | 0.272727 | 0.421053 | 0.333333 | Likely benign | Benign (9 vs 3) | 15 | 11 | 19 | 15 |

**Supplementary Table 3. List of novel heterozygous coding variants identified by exome sequencing and shared by four affected members of the index families.** F1, family 1; F2, family 2.

| Variant (hg19) | Gene | Transcript | Coding impact | Genotype F1 V/7 | Genotype F1 IV/6 | Genotype F1 IV/1 | Genotype F2 III/2 | ACMG interpretation | dbSNP ID | GnomAD MAF |
|---|---|---|---|---|---|---|---|---|---|---|
| chr10: 43595968 A⇒G | *RET* | NM_020630.5 c.135A>G p.A45A | synonymous | Het | Hom | Het | Het | Benign | rs1800858 | 0.731 |
| chr10: 43606687 A⇒G | *RET* | NM_020630.5 c.1296A>G p.A432A | synonymous | Hom | Hom | Hom | Het | Benign | rs1800860 | 0.696 |
| chr10: 43613843 G⇒T | *RET* | NM_020630.5 c.2307G>T p.L769L | synonymous | Hom | Hom | Hom | Hom | Benign | rs1800861 | 0.739 |
| chr10: 43615633 C⇒G | *RET* | NM_020630.5 c.2712C>G p.S904S | synonymous | WT | Het | Het | Ref | Benign | rs1800863 | 0.211 |

**Supplementary Table 4. List of coding *RET* variants identified by exome sequencing in four affected members of the index families.** F1, family 1; F2, family 2; Het, heterozygous; Hom, homozygous; Ref, reference sequence; MAF, minor allele frequency.

| Variant (hg19) | Gene | Transcript | Variant impact | Allelic balance F1 IV/16 | Allelic balance F2 IV/1 | CADD score | DANN score | ACMG interpretation | dbSNP ID | GnomAD MAF | HSF prediction | SpliceAI prediction |
|---|---|---|---|---|---|---|---|---|---|---|---|---|
| Chr4: 42467799 C⇒A | *ATP8A1* | NM_006095.2 c.2325-706G>T | intronic | 0.244 | 0.276 | 0.239 | 0.5351 | Likely benign | NA | 0 | No effect | No effect (Δ score 0) |
| Chr4: 42467877 G⇒A | *ATP8A1* | NM_006095.2 c.2325-784C>T | intronic | 0.485 | 0.304 | 0.169 | 0.4319 | Likely benign | rs1174567821 | 0.0001298 (fails QC) | No effect | No effect (Δ score 0) |
| Chr4: 42467910 G⇒A | *ATP8A1* | NM_006095.2 c.2325-817C>T | intronic | 0.395 | 0.292 | 3.699 | 0.8867 | Likely benign | rs1017292115 | 0.0000969 (fails QC) | No effect | No effect (Δ score 0) |
| Chr4: 42467980 C⇒T | *ATP8A1* | NM_006095.2 c.2325-887G>A | intronic | 0.393 | 0.269 | 0.414 | 0.6193 | Likely benign | rs983021637 | 0.0000652 | No effect | No effect (Δ score 0) |
| Chr4: 46058167 T⇒A | *GABRG1* | NM_173536.4 c.916+2067A>T | intronic | 0.538 | 0.385 | 2.925 | 0.08389 | Likely benign | rs1265993144 | 0 (low coverage, 0.1719 on Bravo) | No effect | No effect (Δ score 0) |

**Supplementary Table 5. List of heterozygous intronic variants identified by genome sequencing in the shared haplotypes and shared by two affected individuals.** F1, family 1; F2, family 2; MAF, minor allele frequency; HSF, human splicing finder; QC, quality control; NA, not available.

| Candidate CNVs | HD-aCGH (F1 IV/2, F1 IV/16, F2 III/2 and F2 IV/1) | Genome sequencing (F1 IV/16, F2 IV/1) | Genes | Segregation in index families | DGV/Clinvar/DECIPHER/gnomAD |
|---|---|---|---|---|---|
| **1** | **Intragenic *SLC30A9* deletion, chr4:~41,997,941 - ~42,038,519 (seen in all four patients)** | **Yes, split reads seen** | ***SLC30A9*** | **Segregates in all affected family members and obligate carriers (Sanger sequencing)** | **No (there are other bigger deletions/duplications covering large regions of chromosome 4)** |
| 2 | Deletion near chr4:42,544,476 (in F2 III/2 and F2 IV/1 only) | Yes, split reads seen | n/a | NA | Yes |
| 3 | Deletion near chr4:46,057,135 (in F1 IV/2, F1 IV/16 and F2 III/2 only) | Yes, homozygous deletion | *GABRG1* | NA | Yes |
| 4 | Deletion near chr4:46,119,808 (in F1 IV/2, F1 IV/16 and F2 III/2 only) | Yes (F1 IV/16 only) | n/a | NA | Yes |

**Supplementary Table 6. Features of the copy number variants (CNVs) identified in the index families by means of high-density CGH array (HD-aCGH) in the shared haplotypes.** F1, family 1; F2, family 2; NA, not applicable. Genomic coordinates are based on hg19.

| Ensembl ID | Gene name | Log2FC | Adjusted *P* value |
|---|---|---|---|
| | *Lymphoblastoid cells* | | |
| ENSG00000154277 | *UCHL1* | -2.532483 | 0.24050404 |
| ENSG00000064042 | *LIMCH1* | -0.3810092 | NA |
| ENSG00000109133 | *TMEM33* | -0.0950763 | 0.87905434 |
| ENSG00000182308 | *DCAF4L1* | -0.6983994 | 0.55242525 |
| ENSG00000014824 | *SLC30A9* | -0.2450105 | 0.34921171 |
| ENSG00000188848 | *BEND4* | 0.09938348 | 0.9455848 |
| **ENSG00000178343** | ***SHISA3*** | **3.57197309** | **6.23E-05** |
| ENSG00000124406 | *ATP8A1* | 0.13819085 | 0.86455435 |
| | *Skin fibroblasts* | | |
| ENSG00000154277 | *UCHL1* | -0.1673577 | 0.73734339 |
| ENSG00000064042 | *LIMCH1* | -0.9826098 | 0.24204817 |
| ENSG00000109133 | *TMEM33* | 0.13914387 | 0.47476835 |
| ENSG00000182308 | *DCAF4L1* | 0.45839199 | 0.78228728 |
| ENSG00000014824 | *SLC30A9* | -0.0263056 | 0.87270386 |
| **ENSG00000178343** | ***SHISA3*** | **3.44918541** | **0.00927341** |
| ENSG00000124406 | *ATP8A1* | 0.79644232 | 0.08586073 |

**Supplementary Table 7. RNA-Seq data in EBV-transformed lymphoblastoid cells and skin fibroblasts obtained from affected patients in the index families.** The table shows expression data of genes located in proximity (+/- 1Mb) of the *SLC30A9* deletion as compared to healthy control-derived cells. Significantly different genes are highlighted in bold. *BEND4* was found to not be expressed in skin fibroblasts. Log2FC, log2 fold change; NA, not applicable.

| Ensembl ID | Gene name | Log2FC (fed) | Adjusted *P* value | Log2FC (starved) | Adjusted *P* value |
|---|---|---|---|---|---|
| ENSG00000128272 | *ATF4* | -0.15785 | 0.344044 | -0.23173 | 0.128399 |
| ENSG00000118217 | *ATF6* | 0.213698 | 0.163117 | 0.353613 | 0.010046 |
| ENSG00000171791 | *BCL2* | -0.13692 | 0.206348 | -0.07103 | 0.559123 |
| ENSG00000127022 | *CANX* | 0.249952 | 0.13152 | 0.453255 | 0.001906 |
| **ENSG00000064601** | ***CTSA*** | **0.533541** | **2.63E-08** | **0.569653** | **2.07E-09** |
| **ENSG00000117984** | ***CTSD*** | **0.63109** | **1.38E-06** | **0.512179** | **0.000143** |
| **ENSG00000135047** | ***CTSL*** | **1.407843** | **5.31E-10** | **1.807082** | **4.83E-16** |
| ENSG00000175197 | *DDIT3* | 0.167375 | 0.567807 | 0.443483 | 0.060316 |
| ENSG00000136986 | *DERL1* | 0.380945 | 3.12E-05 | 0.408283 | 6.35E-06 |
| ENSG00000072849 | *DERL2* | -0.01068 | 0.943291 | -0.07024 | 0.537572 |
| ENSG00000099958 | *DERL3* | 0.310455 | 0.120482 | -0.22166 | 0.289455 |
| ENSG00000134109 | *EDEM1* | 0.175518 | 0.305967 | 0.352517 | 0.01605 |
| **ENSG00000088298** | ***EDEM2*** | **0.871663** | **9.14E-14** | **0.89546** | **1.37E-14** |
| ENSG00000116406 | *EDEM3* | 0.035561 | 0.878271 | 0.14084 | 0.440764 |
| ENSG00000144895 | *EIF2A* | -0.29299 | 0.000569 | -0.23522 | 0.008047 |
| ENSG00000172071 | *EIF2AK3* | -0.03569 | 0.89528 | 0.094847 | 0.679476 |
| ENSG00000178607 | *ERN1* | 0.030204 | 0.840187 | -0.01413 | 0.92379 |
| **ENSG00000166598** | ***HSP90B1*** | **0.494064** | **0.000329** | **0.687324** | **1.45E-07** |
| **ENSG00000044574** | ***HSPA5*** | **0.606692** | **2.24E-06** | **0.692453** | **3.83E-08** |
| **ENSG00000100600** | ***LGMN*** | **0.694667** | **3.59E-09** | **0.780494** | **1.84E-11** |
| **ENSG00000090674** | ***MCOLN1*** | **0.608813** | **1.82E-08** | **0.619615** | **8.83E-09** |
| ENSG00000087074 | *PPP1R15A* | 0.284548 | 0.139955 | 0.449563 | 0.009802 |
| ENSG00000106615 | *RHEB* | 0.191996 | 0.0437 | 0.198238 | 0.036266 |
| ENSG00000071537 | *SEL1L* | 0.180543 | 0.302035 | 0.349324 | 0.020584 |
| ENSG00000162298 | *SYVN1* | 0.056953 | 0.732612 | 0.175494 | 0.185992 |
| ENSG00000100219 | *XBP1* | -0.03313 | 0.91521 | -0.02453 | 0.933593 |

**Supplementary Table 8. RNA-Seq data in CRISPR-edited TT cells.** The table shows expression data of relevant genes in the unfolded protein response, ER-associated degradation, and lysosome pathways (mutant vs control) in both fed and serum starved (3 hours) conditions. Significantly different genes are highlighted in bold. Log2FC, log2 fold change.

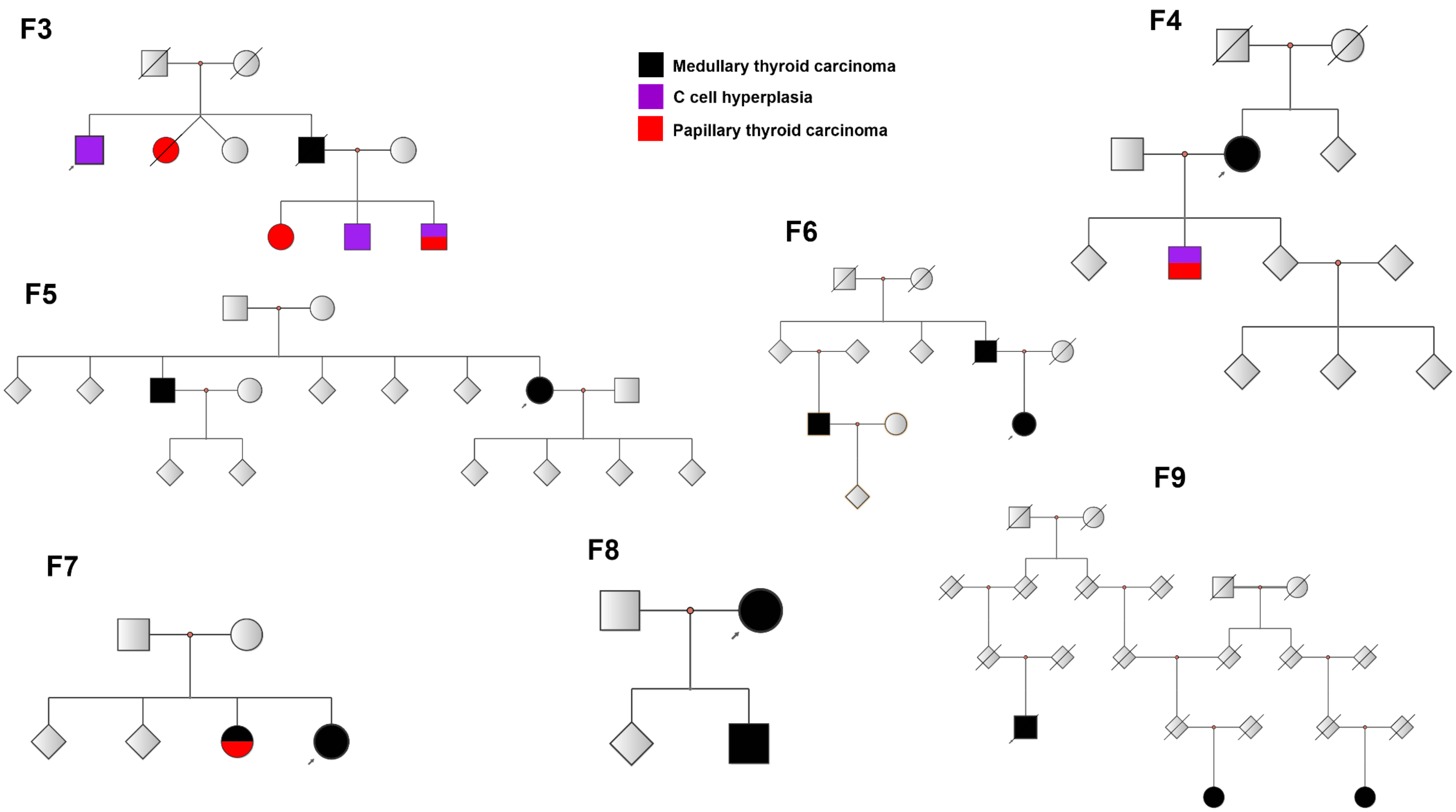

**Supplementary Figure 1. Family trees of the non-index *RET* mutation-negative fMTC families.** Probands are indicated by arrows.

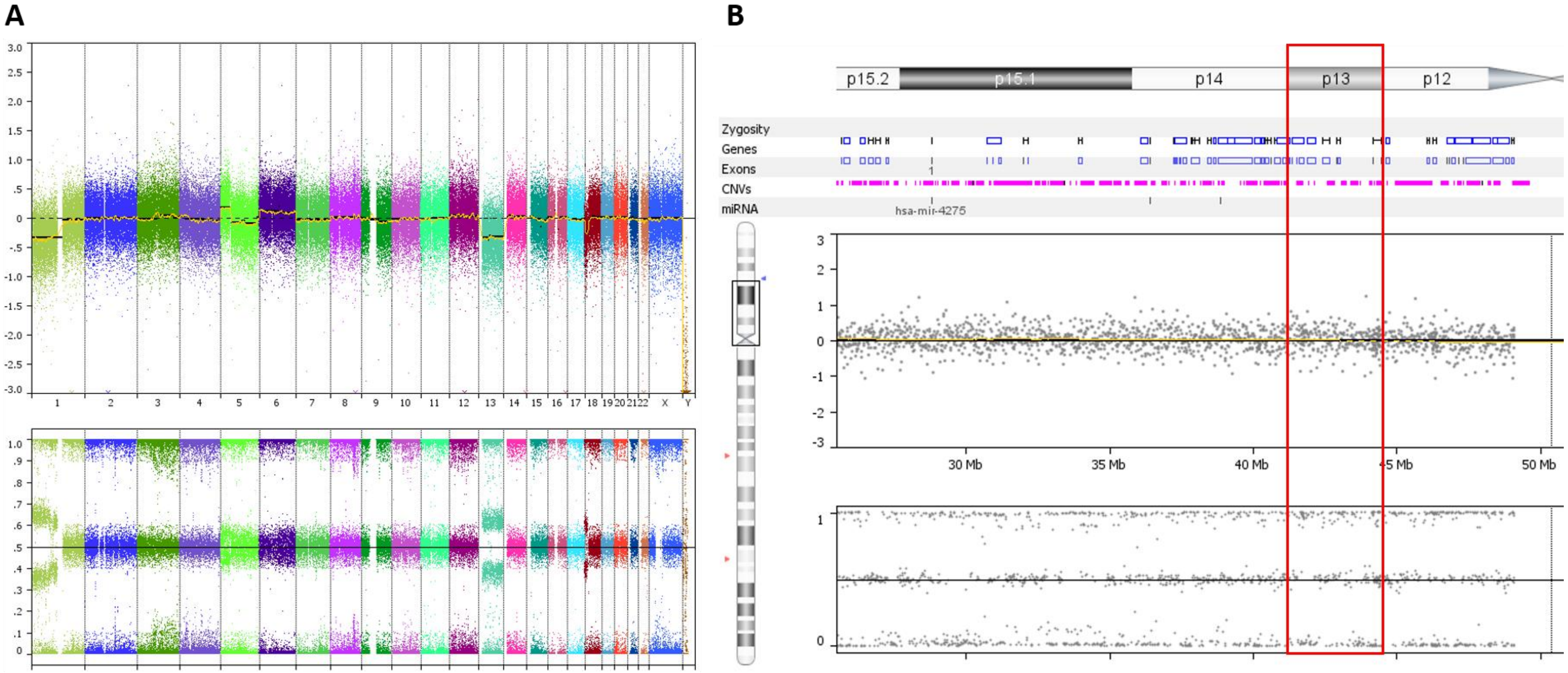

**Supplementary Figure 2. OncoScan assay results in one MTC tumor sample from family 1.** A. Genome-wide view with chromosome number on the x axis and copy number on the y axis expressed as log-2 ratio (top graph) and B allele frequency (bottom graph). Losses at chromosomes 1p, 13, 18p, and a 5p gain are seen. B. Similar graphs as in A showing a closer view of the copy number status on chromosome 4p. The red box highlights chromosome 4p13 with no evidence of somatic gains or losses or loss-of-heterozygosity at this level.

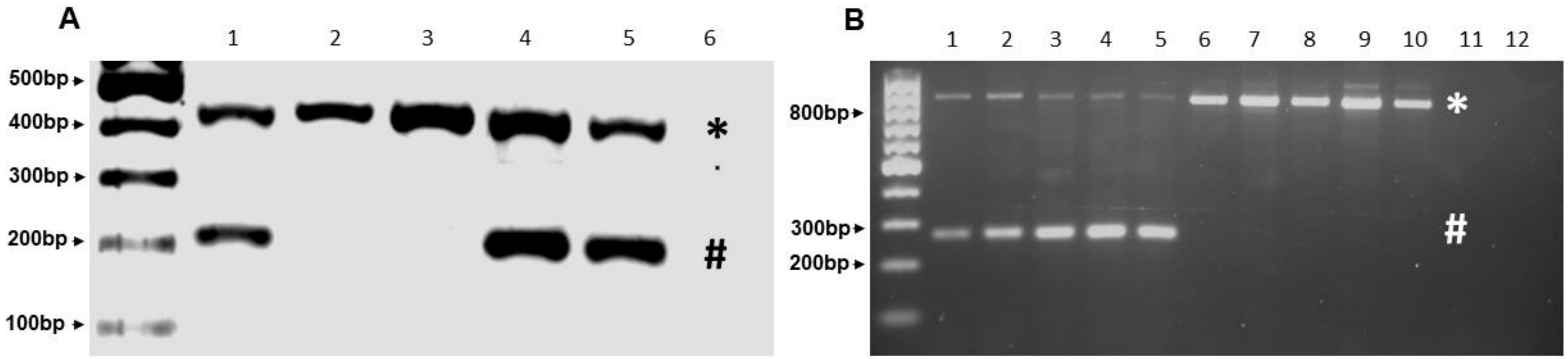

**Supplementary Figure 3. Genotyping and splicing PCR in *SLC30A9* deletion carriers.** A. Example of the duplex PCR employed to determine co-segregation of the *SLC30A9* intragenic deletion in the index families. The asterisk indicates the control allele (*PHOX2B* 3' UTR - 448bp), while the hashtag marks the *SLC30A9* deletion specific allele (217bp). Columns 1, 4, and 5 correspond to patients carrying the deletion, while columns 2 and 3 correspond to unaffected family members. Column 6 shows the no template control. B. Splicing PCR with primers annealing to *SLC30A9* cDNA (exons 1-8). Columns 1-5 correspond to cDNA isolated from peripheral blood leukocytes of affected patients from the index families, while columns 6-10 represent five healthy control samples. The asterisk shows the expected wild type allele (816bp) while the hashtag highlights the mutant allele found exclusively in the deletion carriers (256bp). Sequencing confirmed this fragment to correspond to *SLC30A9* exons 1 and 8. Column 11 shows the -RT control. Column 12 shows the no template control.

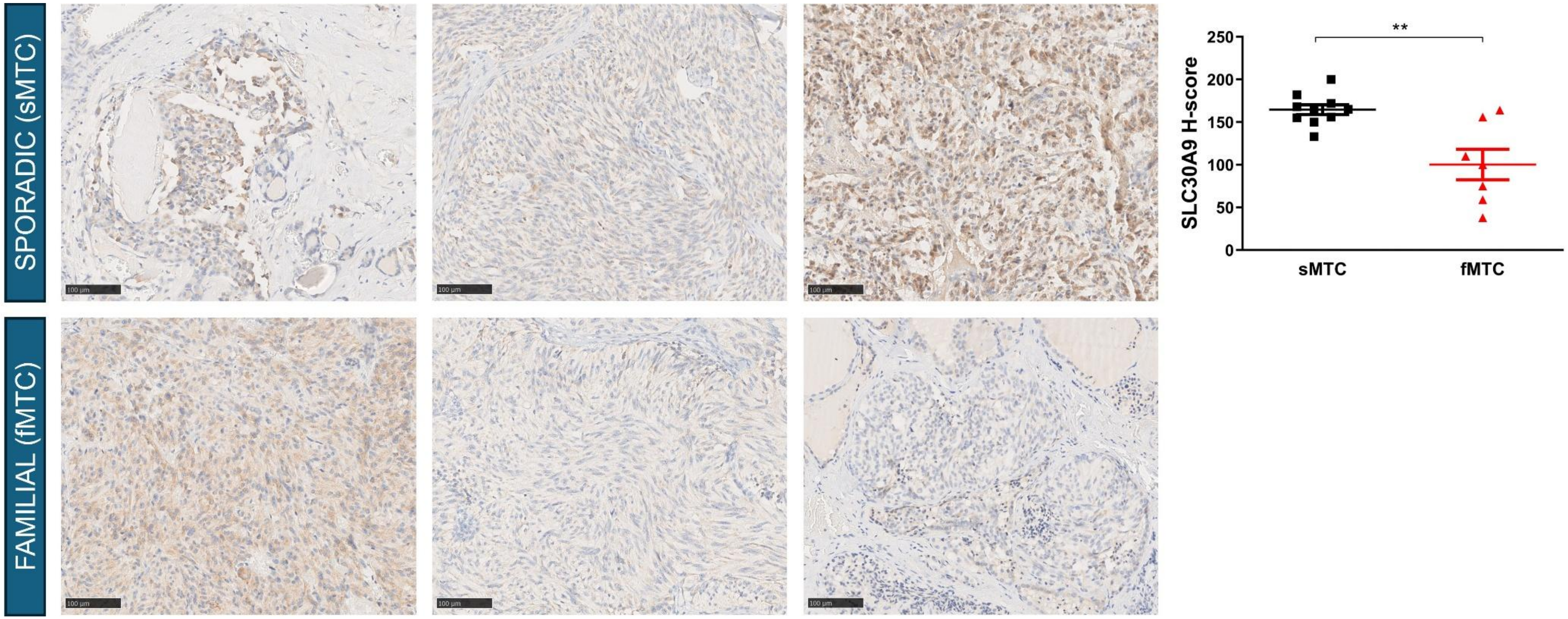

**Supplementary Figure 4. Immunohistochemistry for SLC30A9 in tumor samples from the two families carrying the *SLC30A9* deletion (fMTC) and sporadic medullary thyoid carcinoma controls (sMTC).** Although with some variability among samples, the expression in the familial tumors appeared to be reduced in comparison to sMTC samples (unpaired t test; data shown as individual values, mean, and SEM, n=7-10). Representative images from each of three different cases/group are shown.

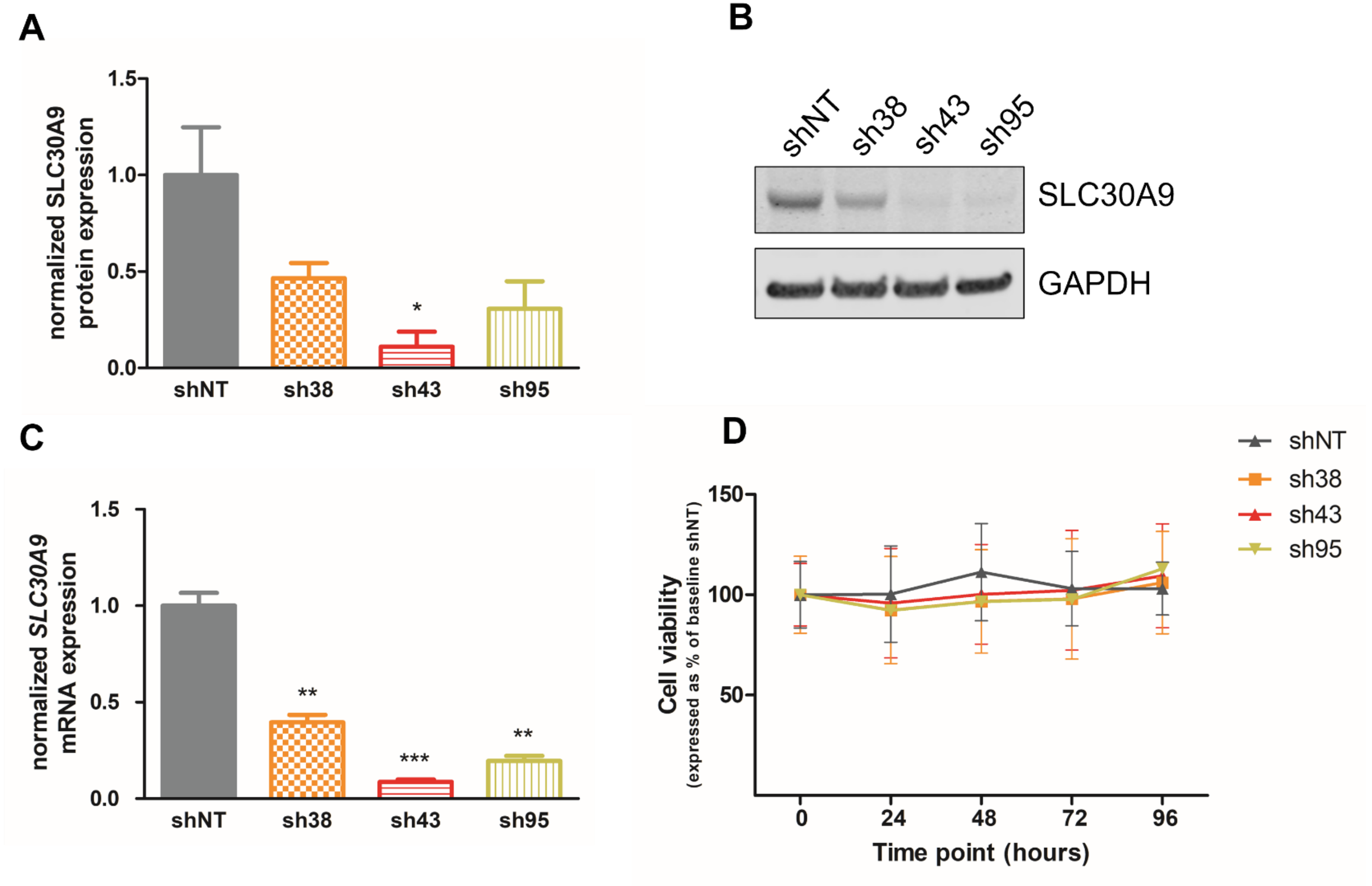

**Supplementary Figure 5. *SLC30A9* silencing in TT cells.** TT cells were transduced with non-targeting (shNT) and *SLC30A9*-specific shRNAs (sh38, sh43, and sh95). Knock-down of SLC30A9 expression was confirmed at both protein (A & B) and mRNA level (C; Kruskal-Wallis with Dunn's post-hoc test; data shown as mean with SEM, n=3). D. No differences were seen in cell viability following *SLC30A9* silencing (Two-Way ANOVA with Sidak's multiple comparisons test; data shown as mean with SEM, n=3).

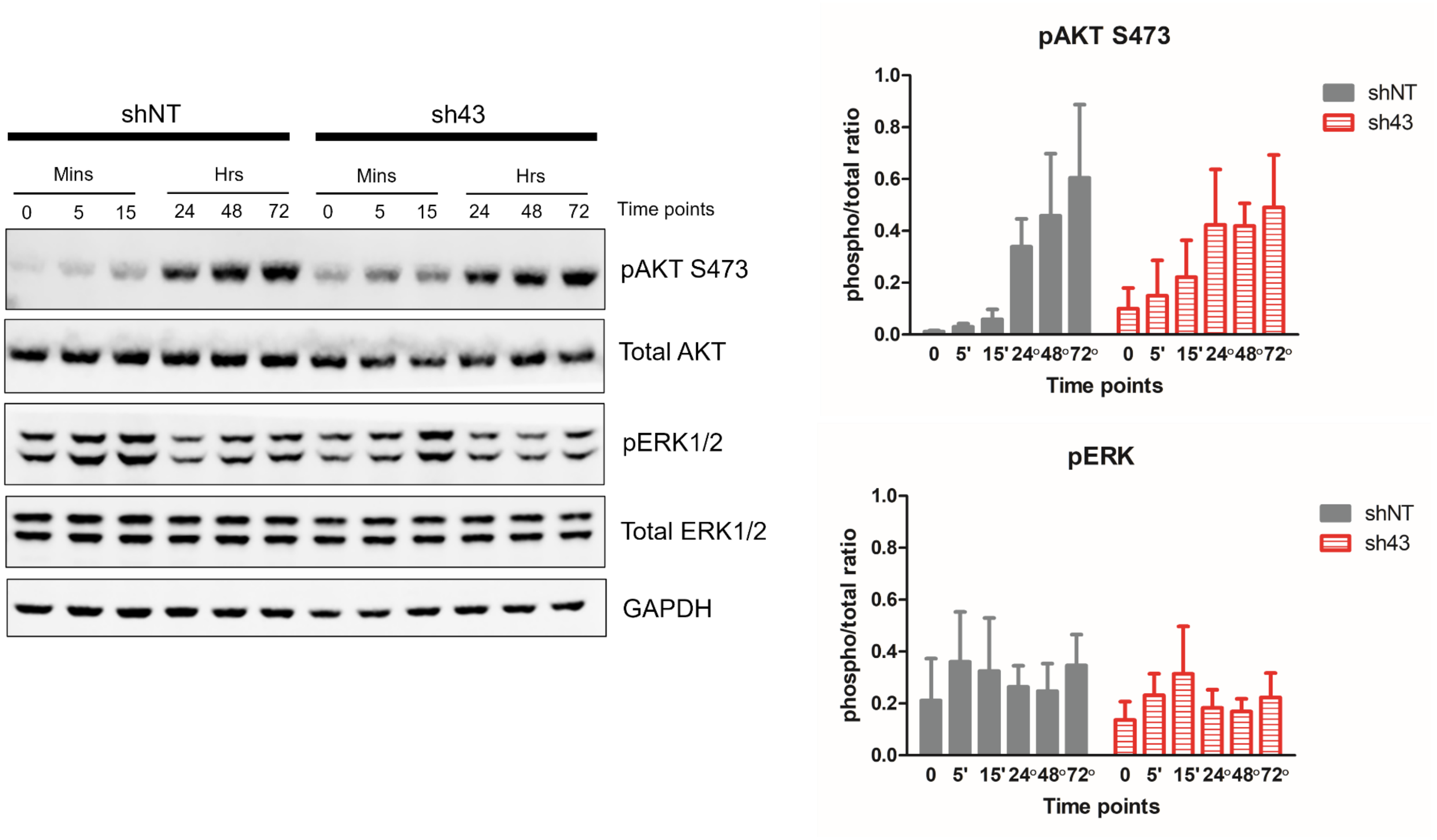

**Supplementary Figure 6. AKT and ERK activation in *SLC30A9*-silenced TT cells.** TT cells transduced with non-targeting (shNT) and *SLC30A9*-specific shRNAs (sh43) were treated with IGF-1 (50nM) for the indicated time points. AKT S473 and ERK phosphorylation were assessed by Western blotting. No consistent differences were observed (Two-Way ANOVA with Bonferroni post-hoc test; data shown as mean with SEM, n=3).

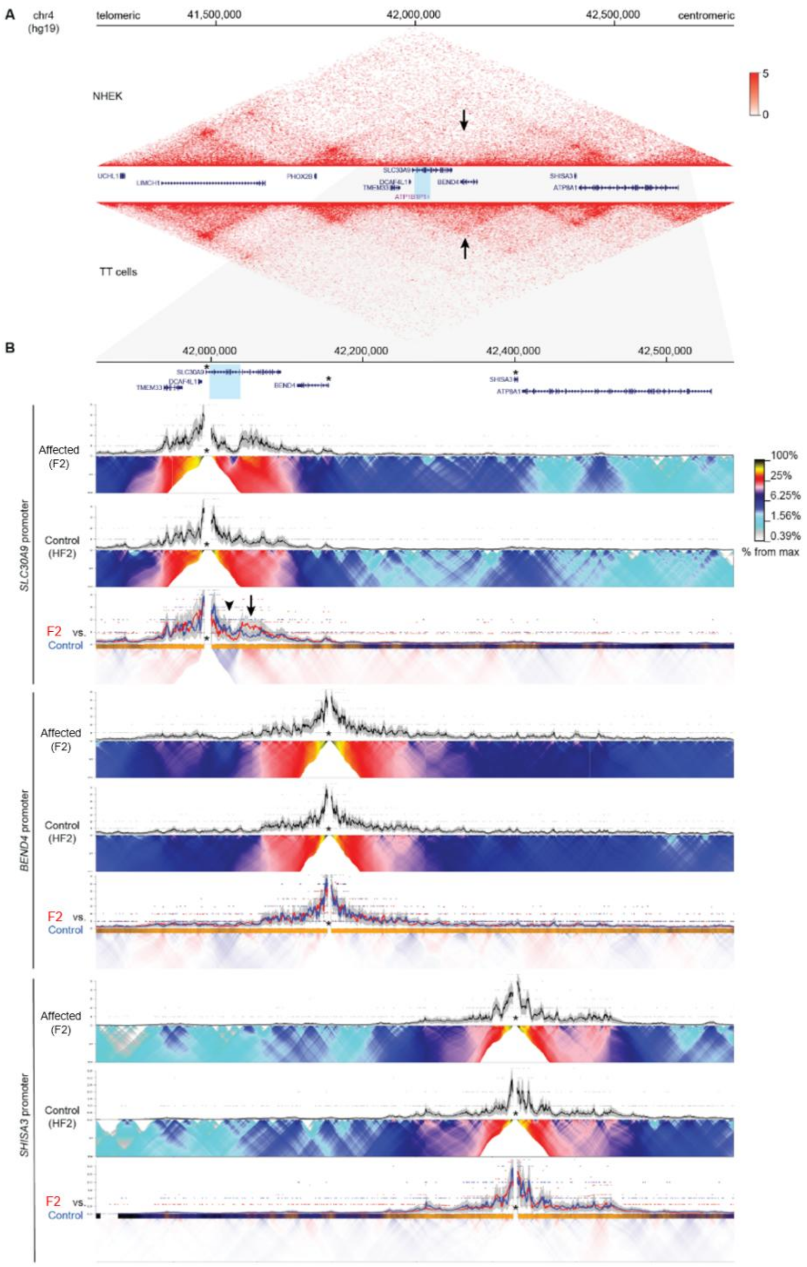

**Supplementary Figure 7. Chromatin interaction analysis at the extended *SLC30A9* locus.**

A. Hi-C data from NHEK cells top; SRR1658691 from [Cell. 2014;159(7):1665-80] and TT cells (bottom; this study) visualized in the UCSC Genome Browser (hg19, chr4:41,200,000-42,800,000), showing KR-normalized contact matrices at 5kb resolution. The deletion within the *SLC30A9* locus is highlighted by blue shading. Notably, the overall TAD configuration is preserved in both NHEK and TT cells. The arrow indicates the position of the TAD containing *SLC30A9*, which is less structured in NHEK cells compared to TT cells.

B. Zoomed-in view of the region (chr4:41,848,255-42,688,255; hg19) and UMI-4C analysis, using the promoters of the *SLC30A9*, *BEND4*, and *SHISA3* genes as viewpoints (indicated by asterisks). UMI-4C contact profiles are shown for F2 and control skin-derived cells. Black lines represent the average normalized UMI counts, with gray shading indicating the standard deviation. Below each contact profile, domainograms (heatmaps) display the mean contact per fragment across a range of window sizes (from 10 to 150 fragments). A comparative analysis overlays the UMI-4C tracks of F2 (red line) and control (blue line) cells, with the interaction profiles scaled relative to each other. The domainogram below highlights regions with enriched contact intensities in F2 (red) and control cells (HF2; blue). Note that the deletion at the *SLC30A9* locus results only in localized changes to chromatin interactions. The *SLC30A9* promoter shows a loss of contacts within the deleted region (arrowhead) and a gain of contacts centromeric to the deleted region (arrow). No changes in long-range interactions were observed for *BEND4*, *SHISA3*, or other nearby genes, including *RBM47*, *UCHL1*, *ATP8A1*, and *GRXCR1* (data not shown).

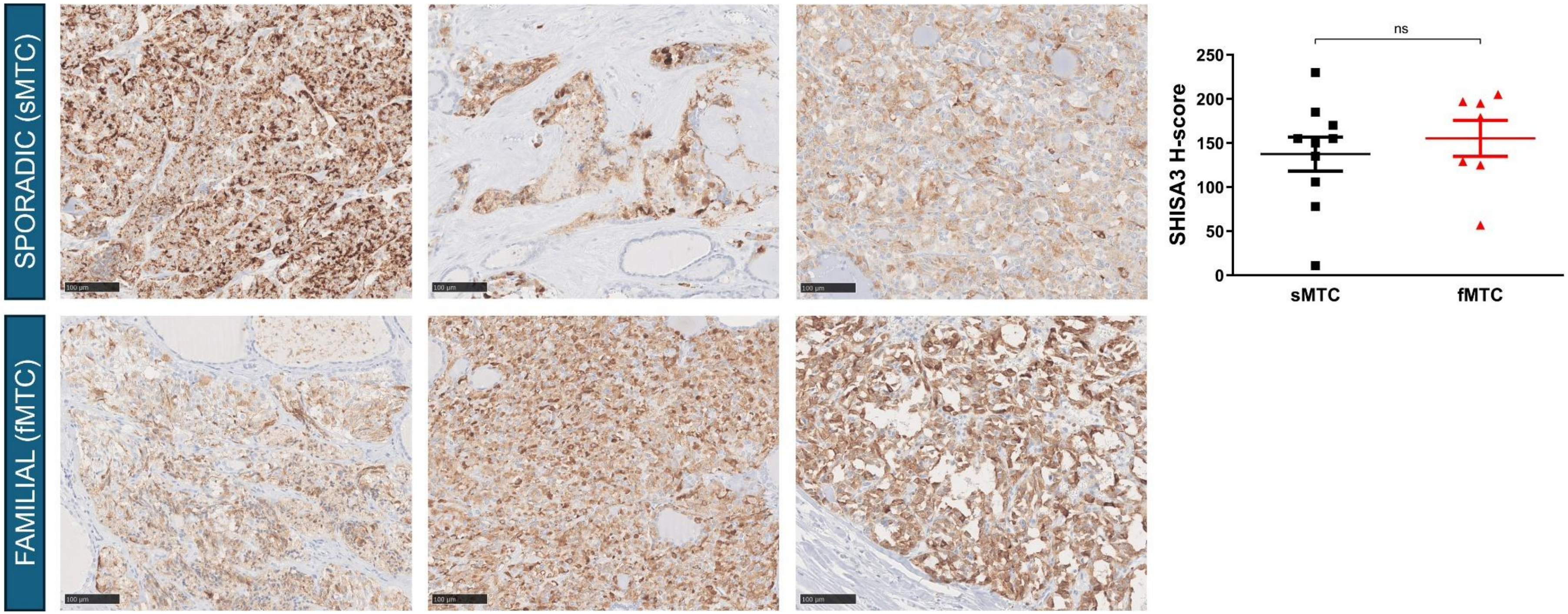

**Supplementary Figure 8. Immunohistochemistry for SHISA3 in tumor samples from the two families carrying the *SLC30A9* deletion (fMTC) and sporadic medullary thyroid carcinoma controls (sMTC).** No consistent differences were seen (unpaired t test; data shown as individual values, mean, and SEM, n=7-10). Representative images from each of three different cases/group are shown.

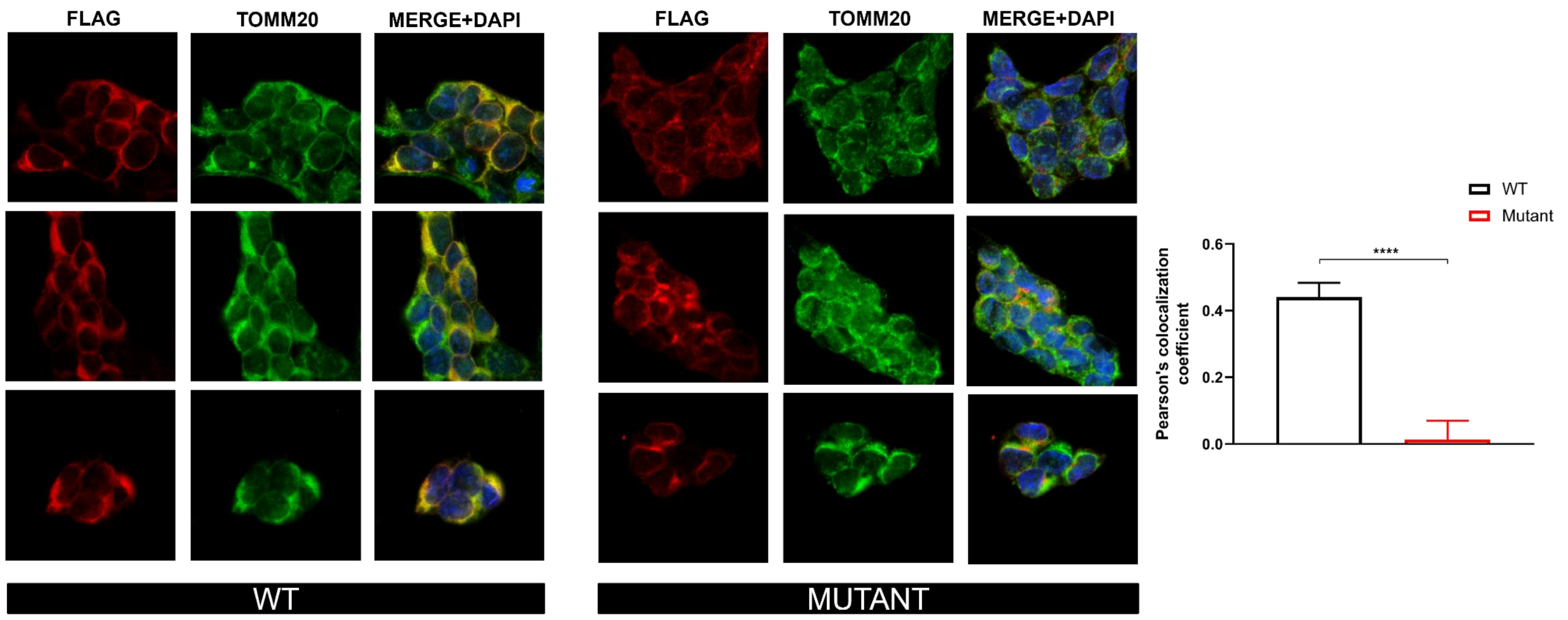

**Supplementary Figure 9. Immunofluorescence shows loss of colocalization of mutant SLC30A9 with TOMM20.** HEK293T cells were transduced with either wild-type (WT) or mutant FLAG-tagged SLC30A9. Full-length (WT) SLC30A9 was found to colocalize with TOMM20, in keeping with mitochondrial localization. However, mutant SLC30A9 showed a perinuclear dot-like expression pattern with loss of colocalization with TOMM20 as assessed with the Pearson's colocalization coefficient (unpaired t test; data shown as mean with SEM). The experiment was repeated twice with at least 100 cells quantified/experiment. Magnification x63.

**Supplementary Methods**

*DNA extractions and RET testing*

Genomic DNA was extracted from peripheral blood leukocytes, saliva, or formalin-fixed paraffin-embedded tumor tissue using commercially available kits (blood: Illustra BACC2 DNA Extraction Kit, GE Healthcare, Little Chalfont, UK; saliva: Oragene-DNA for sample collection and prepIT-L2P for DNA extraction, DNA Genotek, Ontario, Canada; formalin-fixed paraffin-embedded tissue: QIAamp DNA FFPE Tissue Kit, Qiagen, Hilden, Germany). All patients had been tested for the currently recommended germline hotspot *RET* mutations (exons 5, 8, 10, 11, and 13-16) by Sanger sequencing prior to referral apart from seven patients for whom exon 5 had not been sequenced and one patient for whom exons 5 and 8 had not been sequenced. For patients with fMTC, germline *RET* mutations were ruled out prior to inclusion in the study by Sanger sequencing of the whole coding sequence (exons 1-20) of *RET* (NM_020975.6) at the Exeter Genomics Laboratory (Royal Devon and Exeter NHS Foundation Trust, Exeter, UK).

*Multiplex ligation-dependent probe amplification (MLPA)*

A custom MLPA probe mix was designed by MRC Holland (Amsterdam, the Netherlands) to assess *SLC30A9* copy number. The assay contained probes targeting 11 out of the 18 exons of *SLC30A9*, one probe targeting the *ATP1B1P1* pseudogene, 20 reference probes for the purpose of normalization, and three additional control probes to detect experimental problems. Seventy-five ng of genomic DNA were subjected to the MLPA reaction. Data analysis was performed with the Coffalyser.net software (MRC Holland). To rule out the possibility that all tested samples contained the same genetic copy number aberration, the results were first normalized against commercially obtained blood-derived DNA samples of several anonymous male donors (Promega Benelux, Leiden, the Netherlands). Subsequently, the MLPA results were normalized using the population analysis without dedicated reference samples, meaning that the results of each sample were normalized against all other samples.

*Targeted sequencing*

*SLC30A9* (NM_006345.4) and *RET* (NM_020975.6) were sequenced in probands from the *RET* mutation-negative MTC families and all patients with *RET* mutation-negative sporadic MTC using the Fluidigm Access Array (Fluidigm, South San Francisco, CA, USA). One hundred and twenty-five ng of double-stranded DNA were used for each sample. The assay was run on a FC1 Cycler (Fluidigm) according to the manufacturer's instructions. The libraries were sequenced on a MiSeq system (Illumina, San Diego, CA, USA). For data analysis, the sequencing files were aligned to the human genome (GRCh37/hg19) using the Barrow-Wheeler Aligner[1]. Variant calling was performed using Samtools[2] and wAnnovar[3] was used for variant annotation.

*RT-PCR and somatic mutation testing*

In order to assess the effect of the deletion on *SLC30A9* splicing, total RNA was extracted from peripheral blood leukocytes of five patients carrying the deletion (two from family 1 and three from family 2) using the PAXgene Blood RNA Kit (PreAnalytiX GmbH, Hombrechtikon, Switzerland) according to the manufacturer's instructions. Primers for the RT-PCR are available upon request. The mutant band was extracted from the agarose gel using the QIAquick Gel Extraction Kit (Qiagen) according to the manufacturer's instructions.

With the aim of assessing the presence of somatic hotspot *RET* (codons 634, 883, and 918) and *RAS* mutations (codons 12, 13, and 61), DNA was extracted from formalin-fixed paraffin-embedded tumor samples from the index families using the QIAamp DNA FFPE Tissue Kit (Qiagen) and used for subsequent PCR and Sanger sequencing. Primer sequences are available upon request. The PCR reactions were run using *Taq* DNA polymerase (New England Biolabs) on a T100 thermal cycler (Bio-Rad Laboratories, Hercules, CA, USA) and resolved in a 2% agarose gel by electrophoresis. Sanger sequencing was performed under standard conditions by Eurofins (Luxembourg).

*Chromosome conformation capture experiments*

Hi-C was performed in TT cells (2.5 x $10^6$). Nuclei isolation and Hi-C library construction, using the DpnII restriction enzyme, followed a previously established protocol[4] with minor modifications detailed here[5]. The final Hi-C libraries were sequenced using DNBseq technology, producing 100bp paired-end reads, with each sample yielding approximately 400 million raw sequencing read pairs. Hi-C data analysis was performed as previously described with minor modifications[6]. Briefly, Hi-C paired-end reads were mapped to the human genome (GRCh37/hg19) using BWA[1]. Valid ligation events were identified, and PCR duplicates were removed using the pairtools package (https://github.com/mirnylab/pairtools). Unligated fragments and self-ligation events (dangling and extra-dangling ends) were filtered out by discarding paired reads that mapped to the same or adjacent restriction fragments. The filtered pairs file was then converted into a tab-separated values (TSV) file, which served as input for Juicer Tools 1.13.02 Pre to generate multi-resolution Hi-C files[7]. Custom scripts (https://gitlab.com/rdacemel/hic_ctcf-null) were used for the analysis: the hic_pipe.py script first generated TSV files with the filtered pairs, followed by the filt2hic.sh script to produce Juicer Hi-C files. The Hi-C data were visualized in the UCSC Genome Browser (hg19) at a 5kb resolution using the KR normalization option.

UMI-4C library preparation was carried out as previously described[8], with modifications in the 3C library preparation and sequencing library construction, as detailed previously[5]. Experiments were performed in singleton using skin-derived fibroblasts from one affected subject and a control sample, with 1 to 3 x$10^6$ cells as input material. DpnII was used as the restriction enzyme for chromatin fragmentation. The final paired-end sequencing libraries were prepared using the NEBNext High-Fidelity 2× PCR Master Mix (New England Biolabs) and a nested PCR approach, as previously described[8]. Viewpoint primers, comprising upstream (US) and downstream (DS) primers, were designed for DpnII restriction fragments overlapping the transcriptional start site (TSS) of target genes. Both US and DS primers were designed with a melting temperature of 58°C. DS primers were located 5–15bp from the interrogated DpnII restriction site and included the P5-adapter sequence at their 5′ end. US primers were positioned within 100bp of the DpnII restriction site, with minimal overlap with the DS primers. The sequences of the viewpoint primers are available upon request. US or DS primers were pooled for multiplex PCR reactions and used in parallel PCRs with the TruSeq Primer 2.0 (P7 adapter; 5′-to-3′: CAAGCAGAAGACGGCATACGAGAT). Final sequencing libraries were multiplexed and sequenced using DNBseq technology, generating 100bp paired-end reads, with approximately 5–10 million raw sequencing read pairs per viewpoint and sample. For the UMI-4C data analysis, raw fastq files were processed using R version 4.0.3 (2020–10-10) and the R package umi4cpackage 0.0.0.9000 (https://github.com/tanaylab/umi4cpackage)[8]. Contact profiles and domainograms, including difference plots, were generated using the plot function with default parameters and min_win_cov=10, xlim=c(41848255, 42688255).

*Cell viability, proliferation, colony forming, and cell migration assays*

Alamar blue (Thermo Fisher Scientific, Waltham, MA, USA) fluorescent dye was used to measure viability in TT cells following knockdown of *SLC30A9*. Five thousand cells were plated in black multi-well 96 plates in six replicates (Sigma-Aldrich, Burlington, MA, USA) and fluorescence was measured daily for five consecutive days following incubation with the reagent for 3 hours (excitation at 544 nm, emission at 590 nm) using a CLARIOstar Plus Microplate Reader (BMG Labtech, Ortenberg, Germany). For CRISPR-modified TT cells, cell proliferation was assessed by means of live-cell microscopy using an IncuCyte S3 device (Sartorius, Göttingen, Germany). 20 x $10^3$ TT cells were plated in 96-well plates in four replicates and confluency was assessed at intervals of 72 hours for up to 12 days. For colony forming assays, 1.5 x $10^3$ TT cells were plated in 6-well plates in triplicates. After six weeks, cells were fixed with paraformaldehyde and subsequently stained with crystal violet 0.1%. The plates were then scanned using a Li-Cor Odyssey scanner (LI-COR Biosciences, Lincoln, NE, USA). Quantification of colony area and intensity was done using the ImageJ software as previously described[9,10]. For cell migration, 25 x $10^3$ TT were placed within a BioCoat Control Insert (Corning Inc., Corning, NY, USA) in low-serum media (0.1% fetal bovine serum) as duplicates and allowed to migrate against a serum gradient for 24 hours. The migrated cells were then fixed in methanol and stained with 1% Giemsa Blue solution (Sigma-Aldrich). Representative images were taken at 10x magnification from six random fields of the membrane, and average cell count/field was calculated.
